# A standardized instrument quantifying risk factors associated with bi-directional transmission of SARS-CoV-2 and other zoonotic pathogens: The COVID-19 Human-Animal Interactions Survey (CHAIS)

**DOI:** 10.1101/2021.09.21.21263227

**Authors:** Jonathon D. Gass, Kaitlin B. Waite, Nichola Hill, Kathryn Dalton, Kaitlin Sawatzki, Jonathan Runstadler, Meghan F Davis, for the CHAIS Expert Review Group

## Abstract

SARS-CoV-2 (CoV-2), which surfaced in late 2019 in Wuhan City, China, most likely originated in bats and rapidly spread among humans globally, harming and disrupting livelihoods worldwide. Early into the pandemic, reports of CoV-2 diagnoses in pets and other animals emerged, including dogs, cats, farmed mink and some large felids (tigers and lions) from various countries. While most CoV-2 positive animals were confirmed to have been in close contact with CoV-2 positive humans, there has been a paucity of published evidence to-date describing risk factors associated with CoV-2 transmission among humans and domestic and wild animals. The COVID-19 Human-Animal Interactions Survey (CHAIS) was developed through a cross-CEIRS Center collaboration to provide a standardized survey describing human-animal interaction during the pandemic and to evaluate behavioral, spatiotemporal, and biological risk factors associated with bi-directional zoonotic transmission of CoV-2 within households and other shared environments. CHAIS measures four broad domains of transmission risk; 1) intensity and risk of infection among human hosts, 2) spatial characteristics of shared environments, 3) behaviors and human-animal interactions, and 4) animal susceptibility to infection and propensity for onward spread. Following the development of CHAIS, with a One Health approach, a multidisciplinary group of experts (n=20) was invited to review and provide feedback on the survey for content validity. Expert feedback was incorporated into two final survey formats-a long-form and an abridged version for which specific core questions addressing zoonotic and reverse zoonotic transmission were identified. Both forms are modularized, with each section having the capacity to serve as independent instruments, allowing researchers to customize the survey based on context and research-specific needs. Further adaptations for studies seeking to investigate other zoonotic pathogens with similar routes of transmission (i.e. respiratory, direct contact) are also possible. The CHAIS instrument is a standardized human-animal interaction survey developed to provide important data on risk factors that guide transmission of CoV-2 from humans to animals, with great utility in capturing information of zoonotic transmission risk factors for CoV-2 and other similar pathogens.

**Impacts:**

- We present a standardized survey instrument for evaluating risk factors associated with bi-directional zoonotic transmission of SARS-CoV-2 and other similarly transmitted pathogens in household and other settings where humans and animals share close contact.
- The CHAIS instrument evaluates behavioral, spatiotemporal, and host determinants of transmission risk of SARS-CoV-2 and is highly adaptable for use in studies seeking to investigate other zoonotic pathogens such as influenza viruses.
- This standardized instrument will enable pooling of data across studies for meta-analyses to improve predictive models of bi-directional transmission of SARS-CoV-2 among humans and animals and will inform public health prevention and mitigation measures.

## Introduction

SARS-CoV-2 (herein CoV-2), the virus responsible for the COVID-19 pandemic, most likely circulated in bat populations for decades before recombination and transmission to intermediate hosts leading to spillover into human populations, spawning a catastrophic global pandemic and millions of human deaths worldwide (Boni et al., 2020; Johns Hopkins University, 2020; Zhou et al., 2020). Since the initial emergence of the virus, several reports of CoV-2 diagnoses in pets and other animals have been reported. Globally to-date, these reports include infections in domestic dogs, cats, and ferrets, farmed mink, wild mink, captive felids (including puma, cougar, snow leopard, lions, and tigers), and other captive wildlife including gorillas and otters (Hedman, Krawczyk, Helmy, Zhang, & Varga, 2021; OIE, 2020; USDA APHIS, 2020) (Figure 1). Domestic mink infections are particularly important as mink have played crucial roles in multiple disease transmission pathways (including human-to-mustelid, mustelid-to-mustelid, mustelid-to-human, and mustelid-to-feline). Globally, cases have been reported at mink farms in Canada, France, Greece, Latvia, Lithuania, Poland, Netherlands, Spain, Denmark, Italy, the United States, and Sweden. Domestic farmed mink infections have been reported in the thousands, with most reports documenting clinical signs in the animals and some farms reporting unusually high mortality rates (OIE, 2020; RSPCA, 2020). There is also a larger concern of mustelids becoming reservoirs of CoV-2 (Oude Munnink et al., 2021).

**Figure 1:**
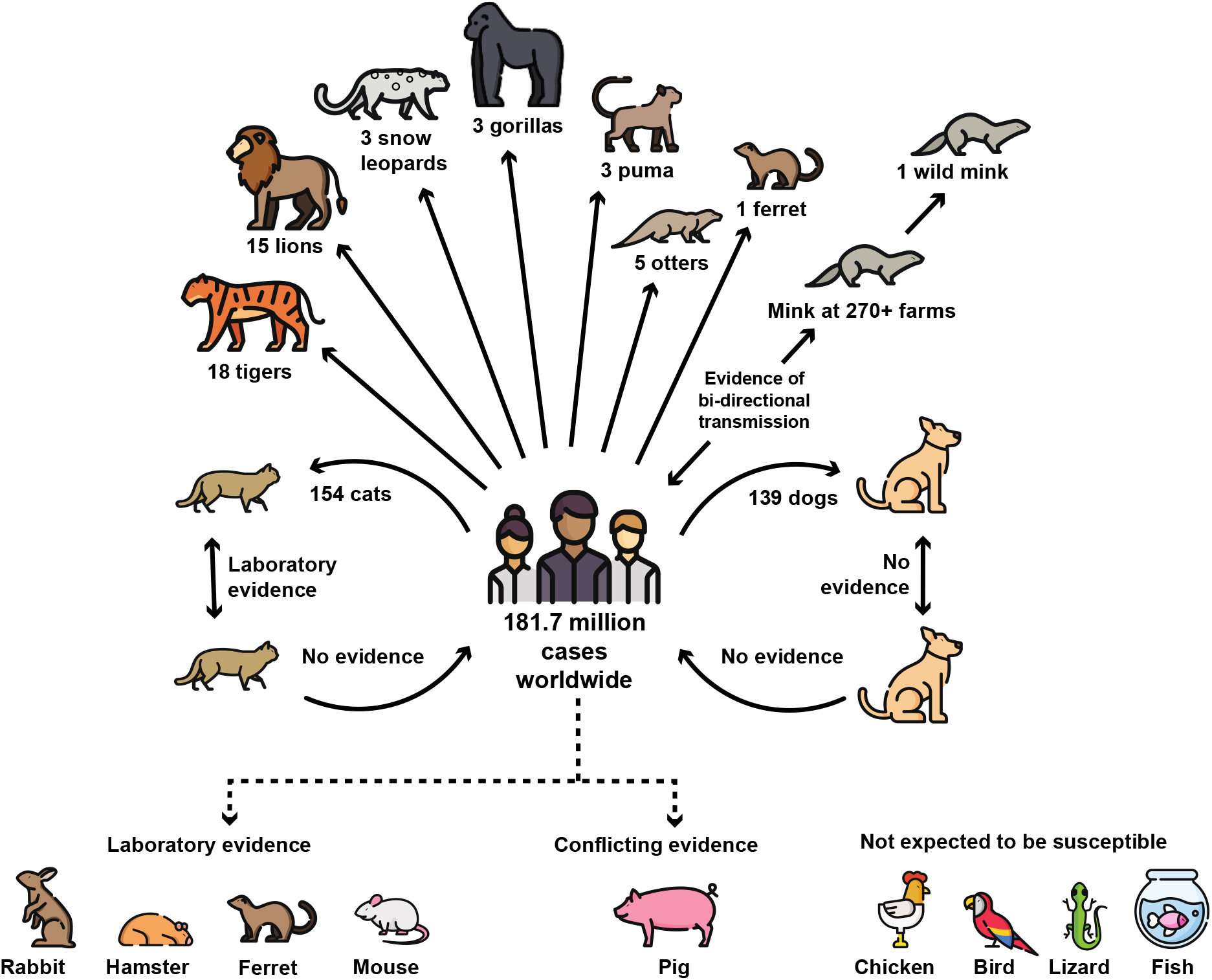
Global evidence to-date of CoV-2 transmission and susceptibility among common household pets and other animals. As of July 29, 2021, there have been 181.7 million human cases of COVID-19 globally (Johns Hopkins University, 2020). Multiple natural cases have been confirmed in animals since January 2020 in 31 countries worldwide. In addition, many CoV-2 outbreaks at mink farms have occurred in the Netherlands, Spain, Denmark, Italy, Sweden, Poland, Latvia, Greece, France, and Canada including secondary transmission from mink back to humans in Denmark (OIE, 2020; Oreshkova et al., 2020; USDA APHIS, 2020). In December 2020, the first free-ranging native wild animal, a wild mink, was confirmed with SARS-CoV-2 near a mink farm in the state of Utah, USA (ProMED, 2020). Laboratory evidence has confirmed that cats are infectious to other cats while there is no evidence of ongoing transmission in dogs (Halfmann et al., 2020). There is no evidence that household pets, including cats and dogs, act as ongoing reservoirs for transmission back to humans. While there have been no natural world cases of CoV-2 in rabbits, hamsters, or mice, laboratory studies have demonstrated these common household animals as susceptible to infection (Liu et al., 2020; Loeffler A, 2011). Multiple studies have concluded that pigs, chickens, other birds, reptiles, and fish are not expected to be susceptible to the virus (Bondad-Reantaso M, 2020; Damas et al., 2020; Luan, Jin, Lu, & Zhang, 2020; Schlottau et al., 2020; Shi et al., 2020). Figure includes modified icons originally made by Freepik from www.flaticon.com.

The probability of interspecies transmission of infectious pathogens is dictated by interactions between human, animal, and environmental dimensions (Plowright et al., 2017). While most CoV-2 positive animals were confirmed to have been in close contact with CoV-2-positive humans in households and other shared environments (Singla et al., 2020), there has been little published evidence to-date demonstrating direct human-to-animal transmission events, nor data describing behavioral, spatiotemporal, and biological risk factors associated with CoV-2 transmission between humans and animals. A deeper understanding of the human-animal interface and potential risk factors associated with CoV-2 transmission between humans and animals is critical for risk prevention and mitigation. While much human-animal interaction data is collected utilizing questionnaires, no standard instrument currently exists for zoonoses. Since CoV-2 testing in animals remains primarily opportunistic, as routine animal testing is not currently recommended (AVMA, 2021; USDA, 2021) research efforts would benefit from the ability to pool data across studies.

Acknowledging the factors that drive interspecies transmission and the need for a standardized tool, the COVID-19 human-animal interactions working group (CHAI-WG) was established to develop the COVID-19 Human-Animal Interactions Survey (CHAIS). The objective of CHAIS is to describe human-animal interactions and evaluate risk factors associated with bi-directional zoonotic transmission of CoV-2 and other similarly transmitted zoonotic pathogens within households and other shared settings. CHAIS evaluates four broad domains of transmission risk: 1) intensity and risk of infection among human hosts, 2) spatial characteristics of shared environments, 3) behaviors and human-animal interactions, and 4) animal susceptibility to infection and propensity for onward spread (Figure 2). In this article, we provide and report on the development of this standard instrument evaluating human-animal interactions in the context of COVID-19, though with broad applicability to multiple zoonotic pathogens, and offer guidance on its many applications in research.

**Figure 2:**
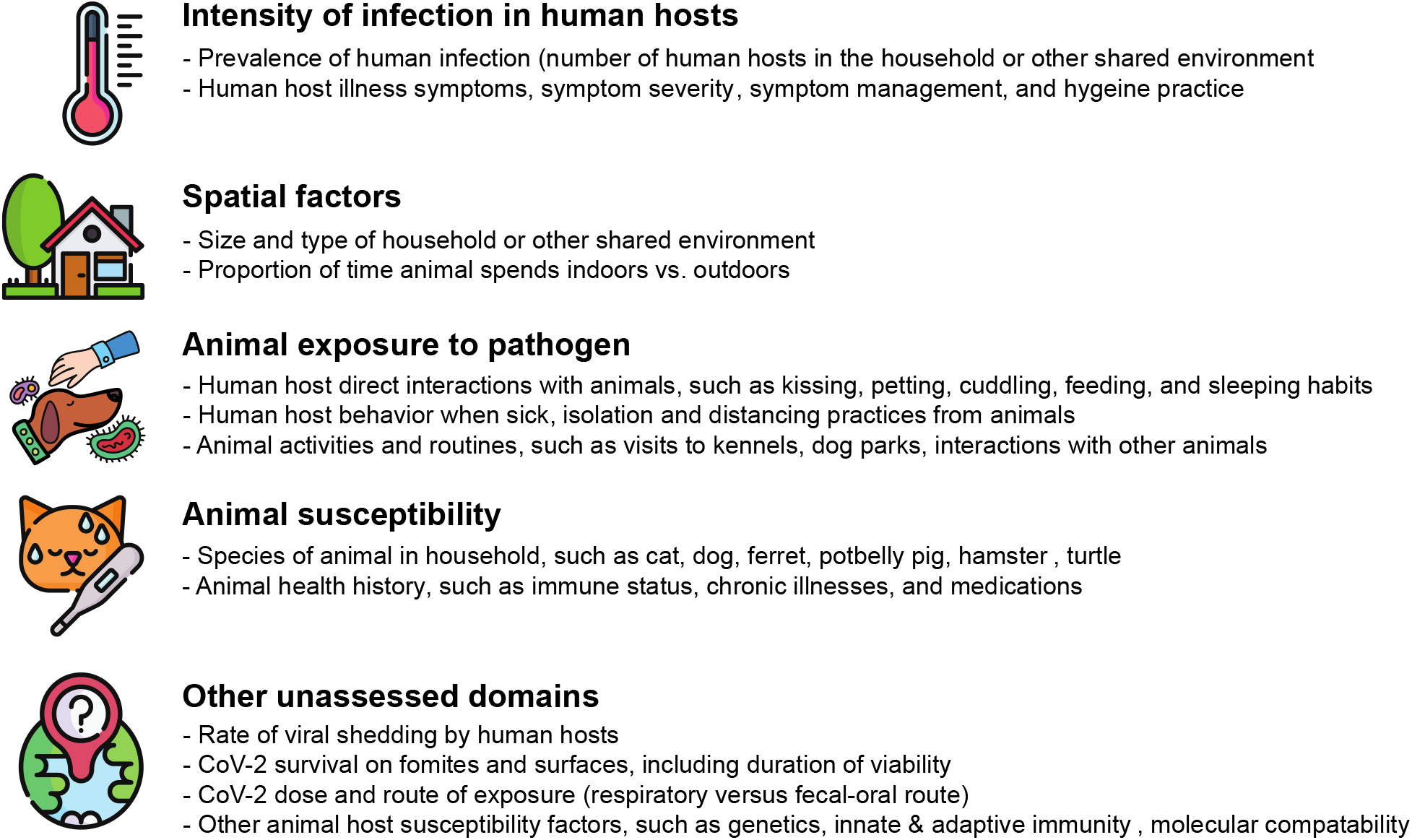
CHAIS domains evaluating CoV-2 transmission from humans to animals in household and other shared environments. The CHAIS instrument focuses on measuring four broad domains which in part are likely to determine reverse zoonotic transmission risk in household settings and other shared environments. Additional domains which the CHAIS instrument does not measure are also described. Figure adapted from Plowright, et al. Pathways to Zoonotic Spillover, Nature, August 2017. Icons made by Freepik from www.flaticon.com.

## Methods

### The COVID-19 human-animal interactions working group (CHAI-WG)

The National Institute of Allergy and Infectious Disease (NIAID) Centers of Excellence for Influenza Research and Surveillance (CEIRS) comprises a network of multidisciplinary collaborating institutions engaged in international surveillance and targeted research on host immune response, viral pathogenesis, emergence, and transmission of influenza viruses. In early 2020, many researchers within the CEIRS network began developing research initiatives in response to the growing number of human cases of CoV-2 globally. Our CEIRS-funded laboratories formed the CHAI-WG to harness both teams’ expertise and expedite the development of a standardized survey instrument given the increasing number of reported cases of domestic and captive animals testing positive for CoV-2 across the globe. The working group comprised faculty and scholars from the fields of veterinary virology, epidemiology, infectious disease ecology, veterinary medicine, and environmental microbiology.

### Development of the CHAIS Instrument

The CHAI-WG collectively determined the structure, types of questions, and intended use for this instrument, including that it be adaptable for use across multiple contexts and for other relevant zoonotic pathogens. Both laboratory teams had previously developed independent instruments before the formation of the CHAI-WG, therefore surveys were combined into a singular preliminary draft. The CHAI-WG refined the combined survey instrument by removing areas of overlap and adding questions based on available and newly published evidence during the evolving pandemic. Additionally, the CHAI-WG identified a comprehensive set of human, animal, and environmental domains that may contribute to transmission pathways for CoV-2 and other similar zoonotic pathogens in the context of close human-animal interaction.

A multi-disciplinary panel of experts (n=20) were invited to critique and provide feedback on the questionnaire for content validation purposes. This expert panel represented multiple areas of discipline including veterinary medicine, human infectious disease, farm-animal and lab-animal medicine, One Health research, virology, microbiology, occupational health, biostatistics, infectious disease epidemiology, veterinary epidemiology, pulmonology, environmental health, human-animal behavioral health, and bioethics. After agreeing to participate, expert reviewers were sent the CHAIS instrument along with a worksheet to complete and return with feedback and edits for each question in the survey, as well as suggestions for additional questions. The goals of this expert-driven pre-testing exercise were to pinpoint problem areas, reduce measurement error and respondent burden, and ensure consistent question interpretation. Edits and feedback from expert reviewers were incorporated into the final survey and any disagreement in these edits was resolved by consensus among CHAI-WG members.

## Results

### The CHAIS Instrument

The CHAIS instrument is offered in two formats, an extended version, E-CHAIS (Appendix 1), and an abridged version, A-CHAIS (Appendix 2), which is comprised of only questions that directly address universal domains associated with zoonotic and reverse zoonotic transmission. Both versions encompass eight modularizable sections that capture multiple levels of human-animal interactions, pet susceptibility to infection, human and animal risk factors and behaviors, and spatiotemporal factors. Each of the eight sections can be used as independent instrument modules and can be adapted to capture other zoonotic and reverse zoonotic pathogens with similar transmission to CoV-2. The questionnaire comprises primarily closed-ended questions with multiple choice responses, as well as follow-up questions based on answers provided by respondents.

Many of the questions in the CHAIS instrument are time-bound to encompass important timepoints in the COVID-19 pandemic, however the dates of time-bound questions may be modified to capture events and behaviors associated with general pathogen transmission or future emergent outbreaks of other zoonoses. We identified January 1, 2020 in this version as a standard point of reference as the start of the COVID-19 pandemic in the United States, allowing us to capture most events retrospectively relating to human-animal interaction during the majority of pandemic timeframe. This date will be especially useful for studies that also include antibody measurement against CoV-2 in humans and/or animals, as historic exposure could have occurred at any time from when human cases emerged in the United States. We identified March 1, 2020 as a general start date on which initial regulatory mandates took into effect, including societal lockdowns in the United States and other countries due to the growing increase in infections. Questions referencing this date will elucidate findings related to behavior modification and/or degree of exposure before and after this date. Several questions also ask whether events occurred, or behaviors were performed in the last six, three, and one-month time period to contextualize responses outside of any calendar-bound period for COVID-19 or other zoonotic disease outbreaks.

Each of the eight sections of the CHAIS instrument is focused on a human subject’s interaction with close-contact animals in environments where humans and animals commonly share contact (Figure 3). Here we provide a brief overview of each section of the CHAIS instrument and how each contributes to accomplishing the aims of this questionnaire.

**Figure 3:**
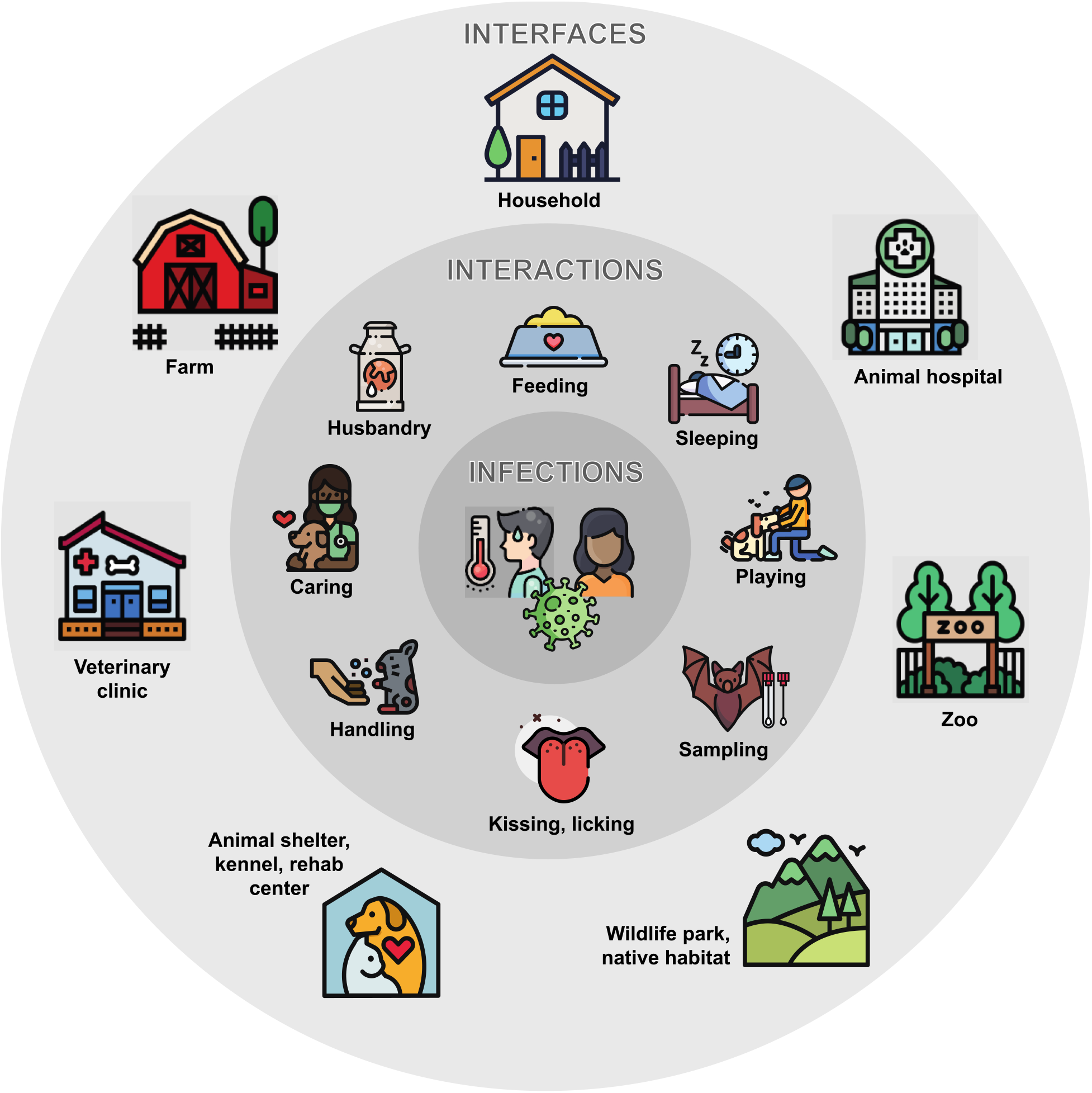
Common interfaces for human-animal interactions associated with reverse zoonotic transmission of respiratory pathogens. The CHAIS instrument focuses on interfaces where animals and humans share close contact, including households, farms, and veterinary settings. Reverse zoonotic transmission events of CoV-2 and other zoonotic pathogens have been postulated to be due to direct interactions between human hosts and animals, specifically kissing, cuddling, playing, feeding, and sleeping habits. Given widespread prevalence of COVID-19 in human populations, the CHAIS instrument seeks to uncover behavioral risk factors and important interfaces for reverse zoonotic transmission events from humans to animals. Not pictured here, the CHAIS instrument also addresses biological and spatiotemporal risk factors underlying these events. Icons made by Iconixar, Photo3idea_studio, Smalllikeart, Pixelmeetup, and Freepik from www.flaticon.com.

The first section, *Household Demographics* [DEM], captures human demographic information for the respondent and each member of the respondent’s household such as date of birth, gender, race, zip code, and employment status. Given each member within a household may uniquely contribute to risk factors of disease transmission to household animals, and vice versa, individual household member demographics allow for attribution throughout the questionnaire. Certain questions in subsequent sections ask the respondent to report events, behaviors, and other data by household member ID. This section also captures information about the household itself, including dwelling type, size in square feet, and number of bedrooms and bathrooms. These questions identify spatial factors that may contribute to exposure to CoV-2 among animals and humans and spread within households of varying type and size. While these spatial factors are isolated to household environments, additional adaptations may be possible for other environments where humans and animals share contact, including zoos, farms, and veterinary settings. In addition, spatial analyses at a broader scale may be possible, including descriptive mapping of participant zip codes with geographically relevant meta-data. Given enough data, spatial statistics may be performed to identify potential infection clustering or quantify spatiotemporal risk factors for infection.

The *Pet Demographics and Behavior* [PetDEM] section compiles baseline information about all animals in the household, such as name, species, age, breed, sex, and duration of ownership. By “pet”, we are referring to an animal, kept primarily for a person’s company or entertainment rather than as livestock or a laboratory animal, however differing definitions for pets across cultures may require further adaptation depending on the context. We capture information about animals that spend significant time outside the home, however the CHAIS instrument is predominantly focused on human-animal interaction within household and other indoor settings. To enable data attribution for individual animals, subsequent sections of the instrument ask that responses are reported individually for each animal in the household. Additional questions identify pet behaviors relative to the household, including duration of time spent outdoors and interactions with animals inside and outside the dwelling. This section also includes questions about whether individual pets are working animals, followed by specific questions inquiring whether they are hunting, protection, detection, service, emotional support, or therapy animals. Service and emotional support animals are often in close contact with mostly one or more members of a household, while therapy animals will also provide therapeutic services to individuals outside of the household, like patients in hospitals and nursing homes, which may contribute to differences in exposure. As working animals have varying roles and risks for disease transmission, the CHAIS instrument asks the participant to describe the nature of working animals’ jobs to capture this wide range of roles. For example, working dogs from the USDA APHIS National Detector Dog program work with United States Customs and Border Protection to help identify prohibited fruit, vegetables, plants, and meat products and are in constant contact with their trainer, as well as travelers, and household members, which may pose unique pathogen transmission risks (McNamara T, 2020; National Detector Dog Training Center, 2021; USCBP, 2021). Further, working dogs hold promise for use in detection for COVID-19 infection in people, activities that could result in exposure to CoV-2 and other pathogens (Hag-Ali et al., 2021; Myers, Hanrahan, Swango, & Nusbaum, 1988; Myers, Nusbaum, Swango, Hanrahan, & Sartin, 1988; Otto et al., 2021).

*The Occupation* [Occ], *Human Travel and Activities* [Travel], and *Animal Worker* [AW] *Sections* include specific questions that help capture the human risk of exposure to CoV-2 through either work, work-related travel, or exposure to household members who have a high risk of occupational exposure. Questions about use of PPE, hand hygiene, and social distancing behaviors are also included. The Occupation Section includes questions about essential worker status and the frequency and duration of time spent working in normal work environments outside of the household. The Human Travel Section includes questions about travel outside of the respondent’s home state and country as well as frequency of activities outside of the home including grocery shopping and attending various types of indoor gatherings. This section also includes questions which identify other areas where there could be significant human-animal interaction and potential interfaces for exposure outside of the home. *The Animal Worker Section* [AW] targets individuals who work with animals in some capacity, primarily those in small and large animal health and husbandry professions. Questions included in this section relate to activities, protective measures, behaviors, and potential exposures that respondents and household members may have while at the workplace.

*The Human Illness History/COVID-19 Section* [HMNill] gathers information about the respondent’s and other household members’ health status and medical history with respect to chronic illnesses and other underlying health conditions that are associated with increased risk of severe COVID-19 illness. This section also captures important information about COVID-19 symptomatology, human-pet interactions, isolation measures, and hand hygiene during periods when respondents or household members were symptomatic with potential COVID-19 illness. Similarly, the *Pet Health History Section* [PetHlth] evaluates illnesses and underlying health conditions that may play a role in the susceptibility and infection severity of pets to contracting COVID-19. This section also serves to measure pet health outcomes and evaluates the odds of these health outcomes relating to human illness in that household.

Building on the other works that have examined zoonoses and risk of pathogen transmission among humans and household pets based on intensity and frequency of human-animal contact (Joosten P, 2020; Morris DO, 2012; Stull JW, 2013), the *Human-Animal Interaction Section* [HAI] captures that intensity and frequency of human-animal contact with more granularity. These human-animal interactions include frequency, duration, and type of contact with pets, such as whether animals sleep in beds with humans in the household and whether they allow pets to kiss or lick their mouth or face. To quantify the closeness of human and animal interactions, and to address the potential for collinearity among variables related to human and animal interactions, the CHAIS instrument includes an interactions index which weighs individual behaviors relative to zoonotic transmission risk (Morris DO, 2012) (Table 1).

**Table 1.**
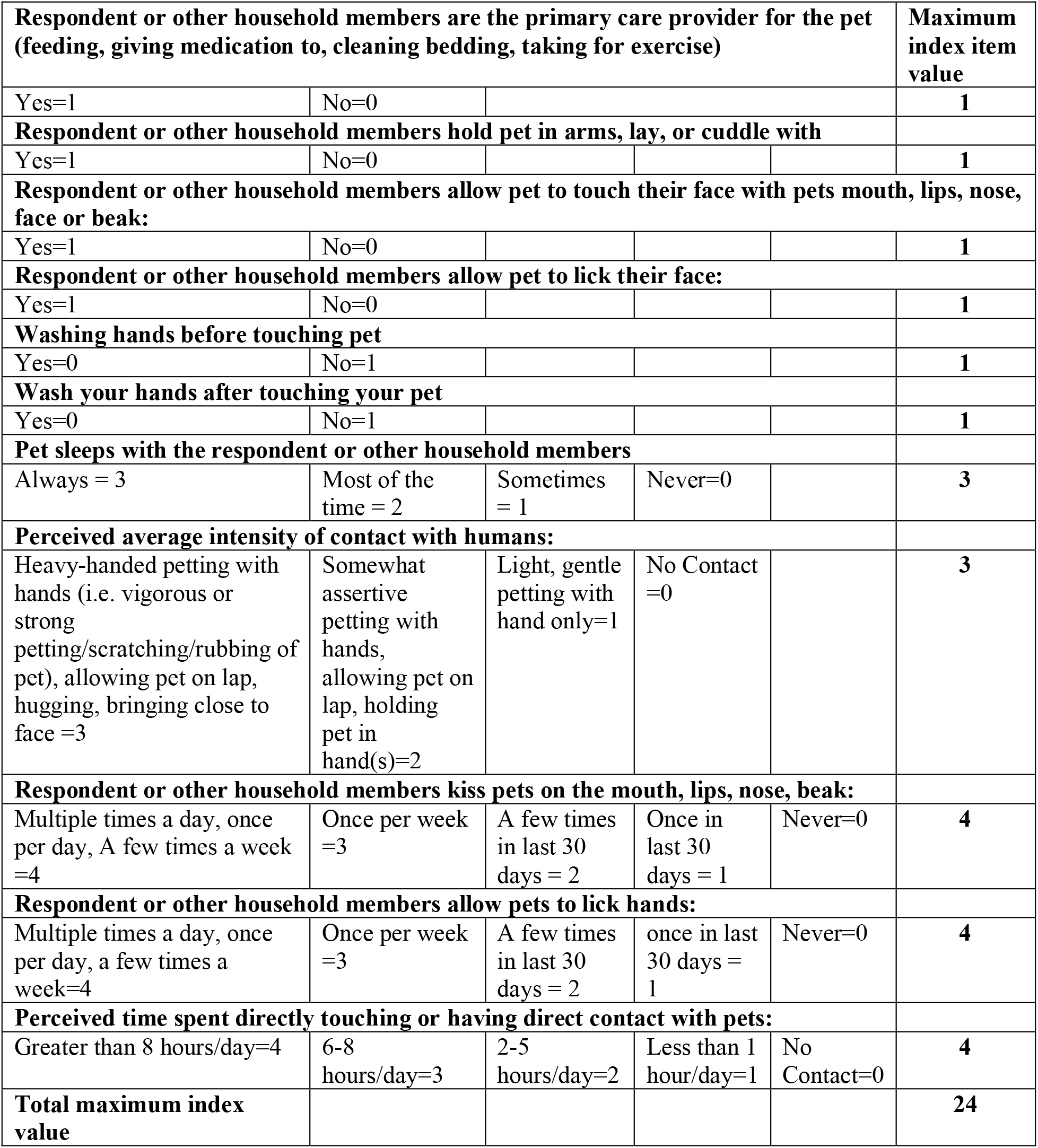
Human-animal interactions closeness index.

## Discussion

The CHAIS instrument serves as a standardized data collection tool to describe human-animal interactions during the COVID-19 pandemic and measure risk factors associated with bi-directional zoonotic transmission of CoV-2 and other similarly transmitted pathogens between humans and animals. Through the measurement of the four broad domains (Figure 2), the CHAIS instrument serves to evaluate behavioral, spatiotemporal, and biological risk factors associated with zoonotic and reverse zoonotic transmission events in household and related settings, and it will facilitate the harmonization of data collection across studies enabling for future data-pooling and meta-analysis of findings. Through proper citation of the CHAIS instrument, cross-study data-pooling and meta-analysis will a) improve our understanding of pathogen exposure and transmission, b) provide a basis for predictive models of bi-directional transmission of SARS-CoV-2 among humans and animals in shared environments, and c) provide an evidence-base for public health guidance. To expand usability, the CHAIS instrument encompasses two versions, an extended (E-CHAIS) version detailing human-animal interactions among participants and household members across multiple pets and animals, and an abridged version (A-CHAIS), which includes core questions addressing zoonotic and reverse zoonotic risk factors for transmission of CoV-2 (Table 2).

**Table 2.** Citation guidance for individual sections for both extended and abridged versions of the CHAIS instrument. The numbers associated with each section represent the question count for that version of CHAIS.

| Sections | Extended CHAIS (E-CHAIS) | Abridged CHAIS (A-CHAIS) |
| --- | --- | --- |
| Household Demographic | DEM-12 | DEM-11 |
| Pet Demographic and Behaviors | PetDEM-13 | PetDEM-6 |
| Occupation Section | Occ-6 | Occ-3 |
| Travel and Activities Section | Travel-10 | Travel-3 |
| Human-Animal Interaction | HAI-11 | HAI-9 |
| Human Illness History/COVID-19 Section | HMNIll-19 | HMNIll-12 |
| Animal Worker Section | AW-7 | AW-4 |
| Pet Health History | PetHlth-27 | PetHlth-13 |
| **Total number of questions** | **105** | **61** |

Both versions of CHAIS contain eight modularizable sections that can be considered individual instrument modules. Individual studies may deploy and use the survey in one of several ways. Researchers may: 1) incorporate either extended or abridged versions of the survey en bloc within their study; 2) select individual modules from either the extended and/or abridged versions of CHAIS and use them independently or in conjunction with another instruments; 3) use individual modules in their entirety with the selection of individual questions from other modules; 4) select individual questions and cite the CHAIS instrument as the source which will allow traceability of the instrument’s usage. While the CHAIS instrument is adaptable, the offering of multiple versions and independent modularizable sections allows for standardization of pet contact instruments when properly cited. Researchers who use either E-CHAIS or A-CHAIS en bloc will simply name one or both instruments with citation, whereas researchers who use a modular approach, such as those already conducting research with need for only certain types of questions covered in specific modules, are encouraged to name the individual modules with citation (Table 3). The CHAIS instrument also allows for minor amendments to identified questions to best serve studies of varying regionality, population, temporality, and cultural diversity (Table 4).

**Table 3.** Citation guidance for modularization and other adaptations for both E-CHAIS and A-CHAIS.

| Example of how indented survey usage | Example citation |
| --- | --- |
| Use of extended CHAIS in its entirety | (E-CHAIS) |
| Use of abridged CHAIS in its entirety | (A-CHAIS) |
| Combination of independent modules used in their entirety | (CHAIS: DEM-12, Occ-2, HAI-9, and AW-4) |
| Use of independent modules in their entirety with isolated questions from other modules within the survey | (CHAIS: DEM-12 + Miscellany) |
| Incorporation of independent questions selected throughout the instrument | (Adapted E-CHAIS) or (Adapted A-CHAIS) |

**Table 4.** Suggested guidance for amending individual questions for differing study contexts. Question numbering depicted below reflect those in E-CHAIS.

|  |
| --- |
| **Regionality Modification Guidance:**<br>CHAIS questions were developed with US regionality in mind and therefore should be amended to better reflect non-US regionality. For example, future studies may need to amend ethnicity response categories to reflect population composition. Studies outside of the US may consider switching to the metric system, including regionally specific invasive and pet animal species, modifying questions to reflect travel within/outside of country location, and providing regionally-relevant examples of pet vaccines and commonly used medicines. Example questions for modification:<br>**Household Demographic questions:** 2,3,6<br>**Pet Demographic question:** 23,24 |
| **Occupation Section question:** 27,29<br>**Travel Section questions:** 32,33,34,35<br>**Pet Health History questions:** 93, 94,95, 96,98 102 |
| **Study Regulatory Modification Guidance:**<br>The CHAIS instrument queries both participant and household member information. Some questions pertaining to household member information might be considered Protected Health Information (PHI) and removal or modification of these questions might be necessary depending on individual research regulatory boards. Example question for modification:<br>**Household Demographic question:** 12 |
| **Timeframe Modifications Guidance:**<br>Time-bound questions have been set to either January 1, 2020 to indicate the start of a post COVID-19 pandemic period in the US or March 1, 2020 to approximate when initial regulatory mandates (shut-down/lockdown events) took effect in most of the United States. These dates may need to be adjusted based on study region and timeframe of such events. Example questions for modification:<br>**Pet Demographic question:** 23,24<br>**Travel section question:** 31, 32, 33, 34, 35, 36ii, 37b ,38 ,39<br>**Human-Animal Interaction questions:** 41, 48, 48 b, 54, 54 b, 55<br>**Human illness History/COVID-19 questions:** 61, 62,67,68<br>**Animal Worker Section:** 80,<br>**Pet Health History questions:** 83, 85,86,97,98,99,103 |
| **Modification Guidance based on future new evidence:**<br>Many questions in this instrument can be adapted and expanded as more information emerges regarding the pathology, symptomology, and other aspects of CoV-2 and COVID-19.<br>**Human illness History/COVID-19 questions:** 60, 62, 63b, 66, 68, 69b |

The CHAIS instrument can be implemented within a broad range of research studies. Studies may include general surveillance of households and other environments where animals and humans share close contact (i.e. farms, veterinary settings, and zoos) regardless of past or present human COVID-19 illness, or studies which specifically target settings where COVID-19 positive humans and/or animals are present. Some examples are described below.

### Research studies that do not include concurrent sampling of humans or animals

The CHAIS instrument can be administered as an epidemiological survey to gather data from community members on human-animal interactions during the COVID-19 pandemic or other zoonotic outbreaks. The CHAIS instrument can rely on self-reported COVID-19 illness history by respondents. Survey questions evaluating human cases of COVID-19 allow for multiple methods for defining a human case. First, respondents are asked to report the date of onset and duration of individual symptoms that are characteristic of COVID-19 illness. Second, respondents are asked whether they a) received a laboratory-confirmed diagnosis, b) were told by a healthcare professional that they had COVID-19, or c) suspected themselves as having the illness but were not tested. Given the latter distinctions, studies that do not include collection of specimens from animals nor humans may rely on respondent self-report of symptomatology among humans and animals. These studies may not confirm but describe potential transmission risks. Finally, aside from assessing risk, many questions on the survey can be used to describe human-animal interactions prior to and during the pandemic, which may facilitate research about the impact of epidemics on human-animal relationships.

### Surveillance and research studies featuring specimen collection from animals or both humans and animals

The CHAIS instrument can contextualize results in studies that include viral and/or antibody testing. For studies in which samples are only collected from animals, the CHAIS instrument may elucidate factors associated with the animal’s exposure. COVID-19 symptoms and illness confirmation questions may provide important self-reported data about human cases of COVID-19. For studies with paired human and animal testing, the CHAIS instrument may be used to measure risk factors associated with zoonotic and reverse zoonotic transmission events. Additionally, studies that include antibody testing of humans and/or animals may use the CHAIS instrument to evaluate transmission risks since January 1, 2020, the estimated origin date of viral circulation in the US to which many questions are time-bound. Researchers should take into consideration the current limitations of antibody testing, including type of immunoglobulin detection, evolving knowledge of immune response and antibody duration in humans and animals, and accuracy of tests, among others (Mathur & Mathur, 2020; Özçürümez et al., 2020; Theel et al., 2020). Researchers also should consider vaccination status of people and animals and are encouraged to add questions as needed in this regard. Finally, studies conducted in households or other environments (such as zoos) where infected humans have similarly reported interactions with multiple animal species, may build upon laboratory animal model studies by elucidating natural-world differences in susceptibility and transmission patterns.

### CHAIS as a standard instrument for other zoonotic pathogens

The CHAIS instrument was specifically designed to be adapted to support studies investigating zoonotic and reverse zoonotic transmission of CoV-2 and similarly transmitted pathogens in settings where animals and humans share close contact, such as zoonotic strains of influenza viruses, *Chlamydophila felis, Bordetella bronchiseptica, Y. pestis, Streptococcus* group A, and methicillin-resistant *Staphylococcus aureus* (MRSA), among others (Davis et al., 2012; Defres, Marwick, & Nathwani, 2009; Loeffler A, 2011; Manian, 2003; Rubinstein, Kollef, & Nathwani, 2008; Shrikrishna D, 2009; Trujillo J, 2012). While there may be important pathological and immunological differences between CoV-2 and other zoonotic pathogens, the CHAIS instrument measures transmission risks which have broad epidemiologic applicability to several host-pathogen systems. This will provide important data on unique transmission pathways for these pathogens. For instance, zoonotic influenza virus transmission is understudied in household environments (Gomaa MR, 2020; Mostafa A, 2018) and the CHAIS instrument may enable an increased understanding of the factors that impact human and animal exposure and infection.

### Limitations

While the CHAIS instrument incorporates spatial characteristics of shared environments and was developed with contributions from experts in environmental health, it alone does not serve as a robust assessment of the built environment and its effects on infectious pathogen spread. We encourage scientists who are interested in investigating the built environment’s role in CoV-2 and other pathogens, to acknowledge the guidelines set forth for environmental assessments and One Health studies, like COHERE (Davis MF, 2017). Such guidelines may inform revision of the CHAIS instrument to accomplish a complete assessment of environmental aspects of zoonotic and reverse zoonotic transmission. For example, to better characterize household environmental risk factors of CoV-2 spread, researchers will want to include questions that capture information about air ventilation systems, sanitary plumbing, building materials, types of home surfaces, and others (Pinter-Wollman N, 2018). These types of additional questions, along with the spatial-characteristic questions already present in the CHAIS instrument will more completely characterize and provide information on the role of environmental factors for pathogen transmission. For studies in which environmental sampling will be collected, including air sampling, expansion of CHAIS could include questions about frequency of sanitation or cleaning of surfaces/shared spaces, cleaning products used, type-of heating and cooling systems used, and other environmental modifiers in the home such as air purifiers or humidifiers. This, with concurrent environmental and human sampling, may provide foundational knowledge of risk factors and environmental drivers of disease transmission. Similarly, the CHAIS instrument is also limited in the depths to which it captures information about wildlife and domestic animal interactions, and wildlife and human interactions. While it provides some framework for these types of questions, for researchers focusing on these interfaces we recommend expanding upon what is provided in the current version of the CHAIS instrument to gather complete information and enhance surveillance efforts of not only CoV-2 but other infectious diseases at wildlife/human/domestic animal interfaces.

The CHAIS instrument is limited when focused on CoV-2 transmission during periods where overlapping seasonal pathogens with similar symptoms to CoV-2 are in circulation (i.e. influenza viruses). Though we believe there is great value in symptom-based reporting for CoV-2 and other diseases (Roland, 2020), the CHAIS instrument is optimally implemented in studies that include molecular or serological testing. In the absence of diagnostics, symptoms due to other respiratory diseases (i.e. influenza viruses) may confound associations between human-animal interactions and zoonotic transmission of CoV-2. For studies that do not include the use of diagnostic confirmatory testing, we recommend that researchers account for co-circulation of known seasonal and endemic pathogens with CoV-2, and acknowledge this when reporting findings.

While this instrument is yet to be fully validated, it was developed building on previously published instruments (Joosten P, 2020; Morris DO, 2012; Stull JW, 2013) and followed an extensive expert review process, satisfying content validation, which can be viewed as the initial step in complete instrument validation (Rattray J, 2007). Given the urgent need to identify risk factors associated with zoonotic and reverse zoonotic transmission of CoV-2 during the COVID-19 pandemic, the CHAI-WG determined that content validation of the instrument was sufficient for public dissemination in anticipation that data collected from multiple studies which adopt CHAIS will inform construct validity of the instrument. This instrument has been populated and is currently available to use through RedCap. To receive a copy of this instrument, contact the corresponding author Dr. Kaitlin B. Waite DVM MPH at. For easy sharing and rapid online deployment, open-access REDCap code for the CHAIS instrument will be publicly available in the future.

Like other large, standardized instruments used across studies, the issue of confounding variables arises. We recommend researchers to use validated instruments where additional questions or adaptations are needed to mitigate these confounding variables. Finally, given the multitude of potential settings and geographies where the CHAIS instrument is deployed, biases in reporting among select populations may occur. Researchers should consider and anticipate these biases when designing research studies including the CHAIS instrument so that this may be accounted for in the analysis.

### Future Directions

The CHAIS instrument was developed to identify factors associated with zoonotic and reverse zoonotic transmission of CoV-2 and other zoonotic pathogens in households and other settings where animals and humans share close contact. Though the CHAIS instrument may be considered an evolving tool to address the multitude of research questions that remain unanswered about exposure risks and transmission dynamics between humans and animals in the COVID-19 pandemic and beyond, we suggest that the research community prioritize the following research questions:

- What are the unique risk factors associated with exposure and transmission at farms, animal hospitals, zoos, shelters, and wildlife rehabilitation centers?
- What are species-specific biologic and behavioral risks for exposure and transmission of zoonotic pathogens?
- What spatial factors are associated with zoonotic and reverse zoonotic transmission of CoV-2 and other similarly transmitted zoonotic pathogens in multiple human and animal shared environments?
- What are the high-risk spillover interfaces of CoV-2 and other zoonotic pathogens between humans and animals, and how might these risks be mitigated?
- As vaccination coverage changes over time, how might human and animal vaccination alter the transmission landscape by adjusting human-animal behaviors and modifying susceptibility?
- Given potential future CoV-2 variants may have enhanced anthropozoonotic potential, which has already been demonstrated by domestic mink to wild mink to human transmission events in the Netherlands (Oude Munnink et al., 2021), what are risk factors associated with future human-to-animal-to-human transmission events?
- What interventions may reduce risks of zoonotic and reverse zoonotic transmission of CoV-2 and other pathogens at interfaces where humans and animals share close contact?

Finally, while a growing number of CoV-2 animal surveillance studies have developed recently, CoV-2 testing in animals remains inconsistent among animal groups (domestic, farmed, and wild), and the importance of active animal surveillance to better understand animal roles as potential reservoirs or intermediary hosts of CoV-2 warrants further investigation (Koopmans, 2020).

## Conclusion

The CHAIS instrument is a standardized tool for evaluating risk factors associated with transmission of CoV-2 and other similarly transmitted pathogens in environments where humans and animals share contact. The CHAIS instrument can be immediately deployed to amplify findings associated with human-animal interactions during the COVID-19 pandemic, which may elucidate risk factors for zoonotic and reverse zoonotic transmission events. Given the gaps in behavioral, spatiotemporal, and biological factors underlying transmission from humans to pets and other animals, the CHAIS instrument has broad applicability, relevance, and importance. As uptake by the scientific community and information about transmission of this virus becomes more robust, we encourage researchers to provide feedback such that this questionnaire may be adapted based on available evidence. Finally, we hope that researchers will cite and provide data for meta-analysis across studies for a more precise understanding of factors associated with zoonotic and reverse zoonotic exposure and transmission of CoV-2 and other zoonotic pathogens.

## Supporting information

Extended CHAIS instrument (E-CHAIS)

Abridged CHAIS instrument (A-CHAIS)

## Data Availability

Authors can confirm that all relevant data are included in the article and/or its supplementary information files.

## Acknowledgements

The authors would like to thank The National Institute of Allergy and Infectious Disease (NIAID) Centers of Excellence for Influenza Research and Surveillance (CEIRS) for providing the opportunity for this collaborative effort. The authors would also like to thank Dr. Cynthia M. Otto for her subject matter expertise. Funding for this project was made possible by NIH/NIAID. The funder had no role in the design, development, nor writing of the CHAIS instrument and manuscript.

## Supplemental Files

- E-CHAIS.pdf - Extended CHAIS instrument (E-CHAIS)
- A-CHAIS.pdf - Abridged CHAIS instrument (A-CHAIS)

