## Extended CHAIS instrument (E-CHAIS) for "A standardized instrument quantifying risk factors associated with bi-directional transmission of SARS-CoV-2 and other zoonotic pathogens: The COVID-19 Human-Animal Interactions Survey (CHAIS)"

| |
|---|
| COVID-19 Study ID: |
| Today's Date |
| Interviewer Initials |

*NOTE: This instrument has been populated and is ready to use through RedCap. Please contact the corresponding author Dr. Kaitlin B. Waite DVM MPH at to receive a copy of this instrument. Open online access through RedCap will be made available in the future. Any text below in red represents code instructions in RedCap.*

### Household Demographics [DEM-12]

**1) What is your date of birth?**

**2) What is your current height?**

**3) What is your current weight?**

**4) What is your gender?**
- ❑ Female
- ❑ Male
- ❑ Transgender
- ❑ Nonbinary
- ❑ Other
- ❑ Do not wish to answer

**5) What is your race?** *Select all that apply*
- ❑ American Indian or Alaska Native
- ❑ Asian
- ❑ Black or African American
- ❑ Native Hawaiian or Other Pacific Islander
- ❑ White
- ❑ Other__________
- ❑ Don't Know
- ❑ Do not wish to answer

**6) What is your ethnicity?**
- ❑ Hispanic or Latino
- ❑ Not Hispanic or Latino
- ❑ Other____________
- ❑ Don't know
- ❑ Not applicable
- ❑ Do not wish to answer

**7) What is your zip code/postal code? ___________**

**8) What type of dwelling do you live in?**

- ❑ Single Family House
- ❑ Townhome
- ❑ Apartment
    - *Branching logic:*
    - How many floors are in your Building?
        - *Dropdown: 2-300, Don't know*
    - Please estimate how many units/apartments are in your Building*:*
        - *Dropdown: 2-300, Don't know*
- ❑ Condominium
    - *Branching logic:*
    - How many floors are in your building?
        - *Dropdown: 2-300, Don't know*
    - Please estimate how many units are in your building*:*
        - *Dropdown: 2-300, Don't know*
- ❑ Other

**9) What is the approximate size of your home in square footage?**

- ❑ Less than 500 square feet
- ❑ 500-1000 square feet
- ❑ 1001-3000 square feet
- ❑ 3001 or more square feet
- ❑ Don't know

**10) How many bedrooms and bathrooms are in your home?** *(if you live in a studio apartment, select "0" for number of bedrooms)*

- Bedrooms ______*{dropdown: 0-20}*__________
- Bathrooms _____*{dropdown: 0-20}*_________

**11) How many people, including yourself, live in your household?** *Please enter the number of household members including yourself.*__________ *{dropdown: 1-20}* ___________

**12) Below, please provide information for up to 8 people currently living with you in your house.**

| Persons ID | Persons Initials | Age in yrs | Sex | Relationship to you |
|---|---|---|---|---|
| *{self-populated hidden field}* | *{free text}* | *{dropdown: <0-100}* | ❑ Female<br>❑ Male<br>❑ Transgender<br>❑ Nonbinary<br>❑ Other: free text | ❑ Mother<br>❑ Father<br>❑ Sibling<br>❑ Child<br>❑ Spouse/Partner |

| | | | | |
|---|---|---|---|---|
| | | | ❑ Do not wish to answer | ❑ Other: |

**Pet Demographics and Behaviors [PetDEM-13]**

**13) Do you have any pet animals currently living *inside* your home?**

- ❑ **Yes**
- ❑ **No**

**13 b) How many total pets do you currently have *inside* your house?**

*By pet, we are referring to an animal, kept primarily for a person's company or entertainment rather than as livestock or a laboratory animal.*

*Note: all animals should be counted individually, except in the event of a group of animals living together in an enclosure, for example, multiple fish in a tank should be grouped and identified as 1 fish tank or multiple hamsters in a cage should be grouped as 1 cage of hamsters.*

- ❑ *{Dropdown scroll 1-50}*

**14) What are your pets NAMES, SPECIES, BREED, SEX, year of birth, and approximate AGE?**

*Please identify up to 10 of the pets currently living **primarily in your house**. Please record your pets in the following order of priority:*

- *1) Companion animal [example cats and dogs]*
- *2) Small companion animals [example rabbits, mink, ferrets]*
- *3) Rodents and Pocket Pet(s) [example: hamsters, gerbils, pet mice, pet rats, guinea pigs]*
- *4) Pet birds [example cockatiel, canaries]*
- *5) Farm animals kept indoors [potbellied pigs, chickens]*
- *6) Other [example water turtles or other reptiles, fish tanks]*

*Note: all animals should be listed individually, except in the event of a group of animals living together in an enclosure, for example multiple fish in a tank should be grouped and identified as 1 fish tank or multiple hamsters in a cage should be grouped as 1 cage of hamsters.*

| Pet Study ID | PETs NAME | Species | BREED | SEX | SEX Alt | Age | How long have you had this pet? |
|---|---|---|---|---|---|---|---|
| *{self-populated* | | | | ☐ Male<br>☐Female | ☐Neutered<br>☐ Spayed | ❑ *0-2yrs* | |

| | | | | | | |
|---|---|---|---|---|---|---|
| *hidden field}* | | | | ☐ **Don't know** | ☐ **Don't know** ☐**N/A** | *{Branching logic:* **Age in months** *{dropdown: 1-24 mo}* ❑ *3-6 yrs* ❑ *7-13yrs* ❑ *14-100yrs* |

*Next, we will ask whether any of your pets are working animals, followed by questions about details of your service animals or therapy animals.*

*Note: Service animals provide routine support for individuals with disabilities or other support needs. Therapy animals provide affection and comfort to people in hospitals, retirement homes, nursing homes, schools, hospices, and disaster areas, among others.*

**15) What is the primary purpose of your pet(s)?**

- ❑ Pet/Companionship
- ❑ Working animal
    - Branching logic: Please select the job of this pet:
        - ❑ Hunting animal
        - ❑ Therapy animal
        - ❑ Emotional support animal
        - ❑ Service animal
        - ❑ Protection
            - Branching logic: Please describe
        - ❑ Detection
            - Branching logic: Please describe
        - ❑ Don't know
        - ❑ Other

**16) You indicated one or more of your pets are considered service or emotional support animals, trained to work for the benefit of an individual with a disability or support needs** *(example: seeing eye dog).* **Please select which pets and describe the services they provide and the frequency of services they provide.**

*{Pet name and ID will be piped-in from the first section}*

1) **Pet Name & Pet ID**
   **Type of service** *Dropdown: emotional support, physical disability or disease support, activities of daily living support, support for mental health diagnosis, Other, Do not wish to answer*
   **Frequency of service** *Dropdown: daily, weekly, monthly, less than monthly*

**17 ) You indicated one or more of your pets are considered therapy animals that provide opportunities for motivation, education, or recreation to enhance the quality of human life**

(*example: therapy dog visiting a nursing home*). **Please select which pets and provide when and where the last therapy session the pet attended.** *Note: If the last therapy session was not in-person but via video conference, please write "Zoom" or "Skype" etc.*

*{Pet name and ID will be piped-in from the first section}*

1) **Pet Name & Pet ID**
   **When_______ _________Where___________________**

**18) Do any pet animals spend time outside the home?** *check all that apply*

☐**Yes** (100% Outdoor pet)
☐ **Yes** *(indoor/outdoor: pet spends 30% of their time in either location)*
☐ **No** *(100% indoor pet except for bathroom breaks and leashed/unleashed accompanied walks)*
☐ **Don't Know**

**19) In the last 30 days, how many hours per day has your pet spent outside?** *check all that apply*

| *{Pet name and ID will be piped-in from first section}* | **100% indoor pet (never outside)** | **Less than 1 hr/ day** | **2-4 hrs /day** | **5-7 hrs/day** | **More than 8 hrs/day** | **100% outdoor pet** |
|---|---|---|---|---|---|---|
| Pet A | ❑ | ❑ | ❑ | ❑ | ❑ | ❑ |
| Pet B | ❑ | ❑ | ❑ | ❑ | ❑ | ❑ |

**20) How would you describe your pet's primary environment in relation to where you live?**
❑ Urban
❑ Suburban
❑ Rural

**21) How much time does your pet spend in their own bed per day?**
*{Pet name and ID will be piped-in from the first section}*
**1) Pet Name & Pet ID**
❑ 1-2hrs
❑ 3-5hrs
❑ 6-8hrs
❑ >8hrs
❑ They do not have their own beds

**22)** *{in multiple pet households}* **What is the frequency of your pets sharing beds with each other?**
❑ Very frequently

- ❑ Somewhat frequently
- ❑ Rare
- ❑ Never, each pet only spends time in their bed
- ❑ They do not have their own beds and do not sleep near each other
- ❑ They do not have their own beds and sleep near each other very frequently, etc.

**23) Do any of your pets currently have direct contact with any of the following animals outside the household?** *Direct contact means touching.*

| *{Pet name and ID will be piped-in from the first section}* | **No Contact** | **Less than once/Month** | **Several times/Month** | **Daily Contact** |
|---|---|---|---|---|
| **a)** Domestic Dog | ❑ | ❑ | ❑ | ❑ |
| **b)** Stray Dog | ❑ | ❑ | ❑ | ❑ |
| **c)** Domestic Cat | ❑ | ❑ | ❑ | ❑ |
| **d)** Feral Cat | ❑ | ❑ | ❑ | ❑ |
| **e)** Wild bird | ❑ | ❑ | ❑ | ❑ |
| **f)** Wild rodent (squirrels, mouse, rat, etc.) | ❑ | ❑ | ❑ | ❑ |
| **g)** Wild Mammals: (bunnies, raccoons, skunks) | ❑ | ❑ | ❑ | ❑ |
| **h)** Horse | ❑ | ❑ | ❑ | ❑ |
| **i)** Pig | ❑ | ❑ | ❑ | ❑ |
| **j)** Cow | ❑ | ❑ | ❑ | ❑ |
| **k)** Chickens or turkeys | ❑ | ❑ | ❑ | ❑ |
| **l)** Sheep | ❑ | ❑ | ❑ | ❑ |

**Other animals (write in): ________________________________________**

*Click here to add another pet that has direct contact with animals outside the home*
*[Another animal contact chart will be made available in RedCap]*

**24) Since January 1st, 2020, have any of your pets had direct contact with any of the following animals inside your home?**

- ❑ Bats
- ❑ Rats
- ❑ Mice
- ❑ Don’t Know

**25) Since January 1st, 2020, have you seen a bat or evidence of a bat inside your home?**

- ❑ Yes
- ❑ No
- ❑ Don’t know

### Occupation Section [Occ-6]

**26) Are you currently employed?**

- ❑ Yes
- ❑ No → *Branching logic: Skip to question 29*

**27) In your occupation, do you work with animals?**

- ❑ Yes
- ❑ No → *Branching logic: Skip to question 27 d*

**27 b) If Yes, what is your occupation?** *Please select all occupation categories that apply to your job***.**

- ❑ Animal breeder
- ❑ Animal trainer
- ❑ Animal control worker
- ❑ Animal scientist or animal researcher
- ❑ Farmer, rancher, farm-worker, or other agricultural worker
- ❑ Non-farm animal caretaker (boarding kennel, doggy daycare, or shelter worker, groomer, etc.)
- ❑ Laboratory animal caretaker
- ❑ Veterinarian
- ❑ Veterinary technologist, technician, or assistant
- ❑ Veterinary student
- ❑ Zoologist or zoo worker
- ❑ Wildlife biologist or wildlife worker
- ❑ Involved in livestock processing for food or fiber production
- ❑ Other_____

**27 c) Please list the most common animals you work with.** *For example: "horses, pigs, sheep, goats"*_______________

**27 d) If No, what is your occupation? _______________**

**28) Are you considered an "essential worker" during the COVID-19 pandemic, as defined by state and local authorities (human health professional, law enforcement, grocery store worker, veterinary medicine professional, etc)?**

- ❑ Yes, I am an essential worker
- ❑ No, I am not an essential worker

**29) Are any of your household members currently employed?**

- ❑ Yes
- ❑ No → *Branching logic: Skip to Travel and Activities Section*

**30) Are any of your household members considered an "essential worker" during the COVID-19 pandemic, as defined by state and local authorities (human health professional, law enforcement, grocery store worker, veterinary medical professional, etc)?**

- ❑ Yes, one or more members of my household are essential workers
  - *Branching logic*: Please identify those household members and their occupation *{Household member Initial will self-populate}*
    Person's Initials_______Occupation:___
- ❑ No, none of my household members are essential workers.

**31) In the last 30 days, if your regular workplace is outside your home, <u>on average</u>, how many hours per week have you worked in your regular workplace?**

- ❑ 0 hours
- ❑ 1-10 hours
- ❑ 11-20 hours
- ❑ 21-30 hours
- ❑ 31-40 hours
- ❑ >40 hours
- ❑ N/A (regular workplace not outside the home)

| Travel and Activities Section [Travel-10] |
|---|

**32) Since January 1st, 2020, in which three towns/cities have you spent the most time? Please write the town name followed by State abbreviation in each field (e.g. Boston, MA)** *(less than 3 just leave blank)*

I have spent the most time in: Town/city name____________________ State_____________
I have spent the second most time in: Town/city name__________________ State_____________
I have spent the third most time in: Town/city name____________________ State_____________

**32 b) Which three towns/cities have you spent the most time within the past:**

**i) 1 month (30 days)**

❑ Same as above

I have spent the most time in: Town/city name____________________ State_____________
I have spent the second most time in: Town/city name__________________ State_____________
I have spent the third most time in: Town/city name____________________ State_____________

**ii) 6 months**

❑ Same as above

I have spent the most time in: Town/city name____________________ State_____________
I have spent the second most time in: Town/city name__________________ State_____________
I have spent the third most time in: Town/city name____________________ State_____________

**33) Since January 1st, 2020, have you traveled to another U.S. state other than the one you live in?**

- ❑ Yes, I have traveled to another U.S. state
  - *Branching logic:* Since January 1st, 2020 how many times have you traveled to another U.S state?
- ❑ No, I have not traveled to another U.S. state
- ❑ Don't Know

**33 b) Did your pet(s) travel with you on any of these trips?**

- ❑ Yes → *Branching logic: Please select which pet*
- ❑ No

**33 c) If yes, please identify the state traveled to, start and end date of travel, and means of transport:**

| State visited | Start Date of travel | End Date of travel | Means of transport |
|---|---|---|---|
| *{Dropdown list of all states}* | | | *select all that apply:*<br>❑ Train<br>❑ Boat<br>❑ Car<br>❑ Bus<br>❑ Airplane<br>❑ Other___ |

**34) Since January 1st, 2020, have you traveled outside of the USA?**

- ❑ Yes, I have traveled outside the USA
  - *Branching logic:* Since January 1st, 2020 how many times have you traveled outside of the USA?
- ❑ No, I have not traveled outside the USA
- ❑ Don't Know

**34 b) Did your pet(s) travel with you on any of these trips?**

- ❑ Yes → *Branching logic: Please select which pet*
- ❑ No

**34 c) If yes, please identify, the country you traveled to, start and end date of travel, and means of transport:**

| Country visited | Start Date of travel | End Date of travel | Means of transport |
|---|---|---|---|
| | | | *select all that apply:*<br>❑ Train<br>❑ Boat<br>❑ Car<br>❑ Bus<br>❑ Airplane |

| | | | |
|---|---|---|---|
| | | | ❑ Other___ |

**35) Since January 1st, 2020, have any household members traveled to another U.S. state other than the one they live in?**

- ❑ Yes, one or more household members have traveled to another U.S. state
- ❑ No household members have traveled to another U.S. state
- ❑ Don't Know

**35 b) If Yes, Did your pet(s) travel with them?**

- ❑ Yes → *Branching logic: Please select which pet*
- ❑ No

**35 c) If Yes, please identify the state they traveled to, start and end date of travel and means of transport:**

| State visited | Start Date of travel | End Date of travel | Means of transport |
|---|---|---|---|
| | | | *(select all that apply):*<br>❑ Train<br>❑ Boat<br>❑ Car<br>❑ Bus<br>❑ Airplane<br>❑ Other___ |

**36) Since January 1st, 2020, have any household members traveled outside of the USA?**

- ❑ Yes, one or more household members have traveled outside the USA
- ❑ No household members have traveled outside the USA
- ❑ Don't Know

**36 b) If Yes, Did your pet(s) travel with them?**

- ❑ Yes →*Branching logic: Please select which pet*
- ❑ No

**36 c) If Yes, please identify, the country they traveled to, start and end date of travel, and means of transport:**

| Country visited | Start Date of travel | End Date of travel | Means of transport |
|---|---|---|---|
| | | | *(select all that apply):*<br>❑ Train<br>❑ Boat<br>❑ Car<br>❑ Bus<br>❑ Airplane<br>❑ Other___ |

**37) How often have you done the following within the past 1 Month (30 days):**
**a) Visited an emergency room for any reason**
☐More than 10 times ☐6-9 times ☐2-5 times ☐1 time ☐Never ☐Don't know
**b) Attended an indoor gathering place like a party, class, concert, or place of worship**
☐More than 10 times ☐6-9 times ☐2-5 times ☐1 time ☐Never ☐Don't know
**c) Visited a primary care doctor for any reason**
☐More than 10 times ☐6-9 times ☐2-5 times ☐1 time ☐Never ☐Don't know
**d) Visited a grocery store, corner store, or pharmacy**
☐More than 10 times ☐6-9 times ☐2-5 times ☐1 time ☐Never ☐Don't know

**ii) How often have you done the following within the past 6 Months:**
**a) Visited an emergency room for any reason**
☐More than 10 times ☐6-9 times ☐2-5 times ☐1 time ☐Never ☐Don't know
**b) Attended an indoor gathering place like a party, class, concert, or place of worship**
☐More than 10 times ☐6-9 times ☐2-5 times ☐1 time ☐Never ☐Don't know
**c) Visited a primary care doctor for any reason**
☐More than 10 times ☐6-9 times ☐2-5 times ☐1 time ☐Never ☐Don't know
**d) Visited a grocery store, corner store, or pharmacy**
☐More than 10 times ☐6-9 times ☐2-5 times ☐1 time ☐Never ☐Don't know

**ii) Since January 1st, 2020, how often have you done the following:**
**a) Visited an emergency room for any reason**
☐More than 10 times ☐6-9 times ☐2-5 times ☐1 time ☐Never ☐Don't know
**b) Attended an indoor gathering place like a party, class, concert, or place of worship**
☐More than 10 times ☐6-9 times ☐2-5 times ☐1 time ☐Never ☐Don't know
**c) Visited a primary care doctor for any reason**
☐More than 10 times ☐6-9 times ☐2-5 times ☐1 time ☐Never ☐Don't know
**d) Visited a grocery store, corner store, or pharmacy**
☐More than 10 times ☐6-9 times ☐2-5 times ☐1 time ☐Never ☐Don't know

**38) Have you been to recreational gatherings with people outside your household (including restaurants) within the past:** *Please be as truthful as possible. Remember: this survey is confidential and this question will help us better understand recreational behaviors during COVID-19* ***(Select all that apply)***

**i) 1 month (30 days)**

a) Indoor: Small, ≤10 people
☐More than 10 times ☐6-9 times ☐2-5 times ☐1 time ☐Never ☐Don't know
b) Indoor: Medium, 11-25 people
☐More than 10 times ☐6-9 times ☐2-5 times ☐1 time ☐Never ☐Don't know
c) Indoor: Large, >25 people
☐More than 10 times ☐6-9 times ☐2-5 times ☐1 time ☐Never ☐Don't know
d) Outdoor: Small, ≤25 people

☐More than 10 times ☐6-9 times ☐2-5 times ☐1 time ☐Never ☐Don't know
e) Outdoor: Medium, 26-100 people
☐More than 10 times ☐6-9 times ☐2-5 times ☐1 time ☐Never ☐Don't know
f) Outdoor: Large, >100 people
☐More than 10 times ☐6-9 times ☐2-5 times ☐1 time ☐Never ☐Don't know
g) I have not been to any recreational gatherings in the last 30 days

**ii) 6 months**

a) Indoor: Small, ≤10 people
☐More than 10 times ☐6-9 times ☐2-5 times ☐1 time ☐Never ☐Don't know
b) Indoor: Medium, 11-25 people
☐More than 10 times ☐6-9 times ☐2-5 times ☐1 time ☐Never ☐Don't know
c) Indoor: Large, >25 people
☐More than 10 times ☐6-9 times ☐2-5 times ☐1 time ☐Never ☐Don't know
d) Outdoor: Small, ≤25 people
☐More than 10 times ☐6-9 times ☐2-5 times ☐1 time ☐Never ☐Don't know
e) Outdoor: Medium, 26-100 people
☐More than 10 times ☐6-9 times ☐2-5 times ☐1 time ☐Never ☐Don't know
f) Outdoor: Large, >100 people
☐More than 10 times ☐6-9 times ☐2-5 times ☐1 time ☐Never ☐Don't know
g) I have not been to any recreational gatherings since in the last 6 months

**38 b) Since March 1, 2020, have you been to recreational gatherings with people outside your household (including restaurants)?** *Please be as truthful as possible. Remember: this survey is confidential and this question will help us better understand recreational behaviors during COVID-19* ***(Select all that apply)***

a) Indoor: Small, ≤10 people
☐More than 10 times ☐6-9 times ☐2-5 times ☐1 time ☐Never ☐Don't know
b) Indoor: Medium, 11-25 people
☐More than 10 times ☐6-9 times ☐2-5 times ☐1 time ☐Never ☐Don't know
c) Indoor: Large, >25 people
☐More than 10 times ☐6-9 times ☐2-5 times ☐1 time ☐Never ☐Don't know
d) Outdoor: Small, ≤25 people
☐More than 10 times ☐6-9 times ☐2-5 times ☐1 time ☐Never ☐Don't know
e) Outdoor: Medium, 26-100 people
☐More than 10 times ☐6-9 times ☐2-5 times ☐1 time ☐Never ☐Don't know
f) Outdoor: Large, >100 people
☐More than 10 times ☐6-9 times ☐2-5 times ☐1 time ☐Never ☐Don't know
g) I have not been to any recreational gatherings since March 1, 2020

**39) Since January 1st, 2020, have you attended daycare, in-person classes, or other schooling outside of the home?**

❑ Yes
❑ No

**40) Since January 1st, 2020, have any members of your household**

**attended daycare, in-person classes, or other schooling outside of the home?**

- ❑ Yes → *Branching logic*: Please identify those household members
- ❑ No

**41) In the last 30 days, while not at work, how often have you worn the following personal protective equipment when outside your home?** *Please be as truthful as possible. Remember: this survey is confidential and this question will help us better understand precautionary behaviors during COVID-19. (select all that apply)*

| | Every time I leave the house | Most of the time | Sometimes | Almost never | Never |
|---|---|---|---|---|---|
| **Disposable gloves like Nitrile or latex gloves** | ❑ | ❑ | ❑ | ❑ | ❑ |
| **Non-disposable gloves like gardening or working gloves** | ❑ | ❑ | ❑ | ❑ | ❑ |
| **Medical grade face masks** | ❑ | ❑ | ❑ | ❑ | ❑ |
| **Other face covering like cloth mask** | ❑ | ❑ | ❑ | ❑ | ❑ |
| **Face shield** | ❑ | ❑ | ❑ | ❑ | ❑ |
| **Other protective wear (please specify)** | | | | | |

**41 b)** *{if medical grade face mask is selected}* **Please describe the type of face mask you wear:**

- ❑ Surgical mask
- ❑ N95 or KN95 mask
- ❑ Respirator
- ❑ Other_____

**Human-Animal Interaction Section [HAI-11]**

**42) Who in your household is most involved in the care of** *{pet name self-populated}*
*{Dropdown list of household members identified}*

| **42 b) Since January 1st 2020, how often has** *{person identified in question 42-self-generated}* **performed the following roles with/for** *{pet name self-populated }* | **Daily** | **Weekly** | **Monthly** | **Never** |
|---|---|---|---|---|
| Fill food and water | ❑ | ❑ | ❑ | ❑ |
| Hold in arms, lay with, or cuddle | ❑ | ❑ | ❑ | ❑ |
| Given medication (when needed) | ❑ | ❑ | ❑ | ❑ |
| Clean their bedding | ❑ | ❑ | ❑ | ❑ |
| Clean their litter *{only appear if cat}* | ❑ | ❑ | ❑ | ❑ |
| Take outdoors for exercise | ❑ | ❑ | ❑ | ❑ |
| Throw a ball, frisbee, or other toys with your dog when playing "fetch" | ❑ | ❑ | ❑ | ❑ |
| Play with toys with your cat *{only appear if cat}* | ❑ | ❑ | ❑ | ❑ |
| Play with toys with this pet | ❑ | ❑ | ❑ | ❑ |

*{Question 42 and 42b will repeat and appear for every pet identified in the beginning of the survey}*

**For the following questions regarding intensity of pet contact, please use the following interaction intensity scale:**

| **Score** | **Interaction Intensity** |
|---|---|
| 0 | No Contact |
| 1 | Light, gentle petting with hand only |
| 2 | Slightly more assertive petting with hands, allowing pet on lap, holding pet in hand(s) |
| 3 | Heavy-handed petting with hands, hugging the pet, bringing pet close to face |

**43) Please describe the intensity, on average, of pet contact from interactions with humans within the household for** *{pet name self-populated}*

| *Pet Name & Pet ID: pipped in form first section* | **0** | **1** | **2** | **3** |
|---|---|---|---|---|
| You *{The participant}* | ☐ | ☐ | ☐ | ☐ |
| Household member {Household members Initials will self-populate} | ☐ | ☐ | ☐ | ☐ |

*{Question 43 will repeat and appear for every pet identified in the beginning of the survey}*

**44) In the last 30 days, have you, or a household member, kissed any of your pets on the mouth, lips, nose, face, or beak?**

- ❑ Yes
- ❑ No
- ❑ Don’t Know

**44 b) In the last 30 days, have you, or a household member, let any pets touch your face, or a household members face, with pets mouth, lips, nose, face, or beak?**

- ❑ Yes
- ❑ No
- ❑ Don’t Know

**44 c) In the last 30 days, has any pet licked you, or a household member, in the face? [**

- ❑ Yes
- ❑ No
- ❑ Don’t Know

**44 d) Which pet(s) have you, or a member in your household, kissed on the mouth, lips, nose, face, beak or been touched or licked on the face by?**
*{Check box select of all pets identified}*

**44 e) Who in your household, and at what frequency, kisses** *{pet name}* **on the mouth, lips, nose, face, beak or lets** *{pet name}* **touch their face with its mouth, lips, face, beak or lets** *{pet name}* **lick their face?**

| | **Multiple times per day** | **Once per day** | **A few times a week** | **Once per week** | **A few times in the last 30 days** | **Once in the last 30 days** | **N/A** |
|---|---|---|---|---|---|---|---|
| You *{The participant}* | ❑ | ❑ | ❑ | ❑ | ❑ | ❑ | ❑ |
| Household member {Household members | ❑ | ❑ | ❑ | ❑ | ❑ | ❑ | ❑ |

| |
|---|
| Initials will self-populate} |

*{44e will appear for every pet selected on 44 d}*

**45) Do you, or any members of your household, allow any of your pets to lick your/their hands?**

- ❑ Yes
- ❑ No
- ❑ Don't Know

**45 b) Which pet(s) lick your, or a household member, hands?** *Select all that apply*
*{Check box select of all pets identified}*

**45 c) Who in your household and at what frequency does** *{pet name}* **lick your, or a household members hands?**

| | Multiple times per day | Once per day | A few times a week | Once per week | A few times in the last 30 days | Once in the last 30 days | N/A | Don't know |
|---|---|---|---|---|---|---|---|---|
| You *{The participant}* | ❑ | ❑ | ❑ | ❑ | ❑ | ❑ | ❑ | ❑ |
| Household member {Household members Initials will self-populate} | ❑ | ❑ | ❑ | ❑ | ❑ | ❑ | ❑ | ❑ |

*{45c will appear for every pet selected on 45b}*

**46) Do any of your pets regularly sleep with you or any household members?** *(for example, on the bed, or on the couch, or in a chair)*

- ❑ Yes: → *Branching logic: Skip to question 46b*
- ❑ No
- ❑ Don't Know

**46 b) Which pets regularly sleep with you or any household members?** *Select all that apply*
*{Check box select of all pets identified}*

**46 c) Who in your household does** *{pet name}* **sleep with, and at what frequency?**

| | Always | Most of the time | Sometimes | Never |
|---|---|---|---|---|

| | | | | |
|---|---|---|---|---|
| You *{The participant}* | ❑ | ❑ | ❑ | ❑ |
| Household member {Household members Initials will self-populate} | ❑ | ❑ | ❑ | ❑ |

*{46c will appear for every pet selected on 46b}*

**47) Excluding any time spent sleeping with a *{pet name}*, how many hours would you estimate *{pet name}* spends sharing the same space, room or being in close proximity to you, or a household member, without physical contact with you?** *(e.g. sitting together on the couch or bed, pet laying at your feet, pet watching you cook or work)*

| | N/A | Less than 1 hour/day | 2-5 hours/day | 6-8 hours/day | Greater than 8 hours/day |
|---|---|---|---|---|---|
| **a)** You *{The participant}* | ☐ | ☐ | ☐ | ☐ | ☐ |
| **b)** Household member {Household members Initials will self-populate} | ☐ | ☐ | ☐ | ☐ | ☐ |

*{47 will appear for every pet identified}*

**48) Excluding any time spent sleeping with a pet, how much time do you, or a household member, spend directly touching or having direct contact with any of your pets?** *(e.g. cuddling on the couch, laying on a human's lap, playing indoors etc.)*

| *Pet Name & Pet ID: pipped in form first section* | N/A | No Contact | Less than 1 hour/day | 2-5 hours/day | 6-8 hours/day | Greater than 8 hours/day |
|---|---|---|---|---|---|---|
| **a)** The [participant] | ☐ | ☐ | ☐ | ☐ | ☐ | ❑ |
| **b)** Household member {Household members Initials will self-populate} | ☐ | ☐ | ☐ | ☐ | ☐ | ❑ |

**Notes on human contact:** ________________________________________

*{48 will appear for every pet identified}*

**49) Since January 1st 2020, Do you wash your hands BEFORE touching your pet**?

| *Pet Name & Pet ID: pipped in form first section* | **Since January 1st, 2020, Do you wash your hands BEFORE touching your pet?** |
|---|---|
| *{pet_name_1}* | ❑ Always<br>❑ Most of the time<br>❑ Sometimes<br>❑ Never<br>❑ Don't Know |
| *{pet_name_2}* | ❑ Always<br>❑ Most of the time<br>❑ Sometimes<br>❑ Never<br>❑ Don't Know |
| *{pet_name_3}* | ❑ Always<br>❑ Most of the time<br>❑ Sometimes<br>❑ Never<br>❑ Don't Know |

**49 b) Since January 1st, 2020, Do you wash your hands AFTER touching your pet?**

| *Pet Name & Pet ID: pipped in form first section* | **Since January 1st, 2020, Do you wash your hands AFTER touching your pet?** |
|---|---|
| *{pet_name_1}* | ❑ Always<br>❑ Most of the time<br>❑ Sometimes<br>❑ Never<br>❑ Don't Know |
| *{pet_name_2}* | ❑ Always<br>❑ Most of the time<br>❑ Sometimes<br>❑ Never<br>❑ Don't Know |
| *{pet_name_3}* | ❑ Always<br>❑ Most of the time<br>❑ Sometimes<br>❑ Never<br>❑ Don't Know |

### General Farm Animal Contact

**50) Since January 1st, 2020, have you had routine close contact with farm animals?**

- ❑ Yes →*will be shown question block for animal professionals*
- ❑ No
- ❑ Don't Know

**50 b) If yes, how would you best describe your interaction with farm animals?** *Select all that apply*

- ❑ I work in the veterinary medical field and treat farm animals
- ❑ I work on a farm or own farm animals
- ❑ I have other routine interactions with farm animals (please describe)_____________

**51) Since January 1st, 2020, how often have you had close routine contact with each of the species listed below?**

| | Multiple times per day | Once per day | A few times a week | Once a week | A few times a month | Once a month | A few times within the 6 months |
|---|---|---|---|---|---|---|---|
| Dairy Cows | ❑ | ❑ | ❑ | ❑ | ❑ | ❑ | ❑ |
| Beef Cattle | ❑ | ❑ | ❑ | ❑ | ❑ | ❑ | ❑ |
| Swine or pigs | ❑ | ❑ | ❑ | ❑ | ❑ | ❑ | ❑ |
| Goats | ❑ | ❑ | ❑ | ❑ | ❑ | ❑ | ❑ |
| Chickens | ❑ | ❑ | ❑ | ❑ | ❑ | ❑ | ❑ |
| Horses | ❑ | ❑ | ❑ | ❑ | ❑ | ❑ | ❑ |
| Sheep | ❑ | ❑ | ❑ | ❑ | ❑ | ❑ | ❑ |
| Other | ❑ | ❑ | ❑ | ❑ | ❑ | ❑ | ❑ |

**51 b) In your own words, please describe the type of contact you routinely have with each of the farm animals you listed daily/weekly/monthly interaction within question #56. (For example:** *I feed 12 backyard chickens daily and I clean a horse barn housing 3 horses weekly; or I perform surgery on livestock as part of my veterinary practice, etc.*)

---

*{this question will only appear if selected yes to question #50}*
**52) You noted interaction with farm animals above. Please answer the following questions about livestock interactions with dogs, cats, wildlife, and other animals.**

| | Always | Most of the time | Sometimes | Never | I don't know |
|---|---|---|---|---|---|

| | | | | | |
|---|---|---|---|---|---|
| **Have you ever seen wildlife in your feed? ( e.g. birds, rodents)?** | ❑ | ❑ | ❑ | ❑ | ❑ |
| **Have you ever seen wildlife in your barn/pasture?** | ❑ | ❑ | ❑ | ❑ | ❑ |
| **Have you ever seen dogs and cats in your grain storage, or in your grain?** | ❑ | ❑ | ❑ | ❑ | ❑ |
| **Have you ever seen cats in your barn and/or pasture areas?** | ❑ | ❑ | ❑ | ❑ | ❑ |
| **Have you ever seen dogs access barns and/or pasture areas** | ❑ | ❑ | ❑ | ❑ | ❑ |
| **Are rodent entryways and denning places in buildings eliminated if found** | ❑ | ❑ | ❑ | ❑ | ❑ |
| **Do you implement an integrated pest management program** | ❑ | ❑ | ❑ | ❑ | ❑ |
| **Have you ever (or since January 2020) seen bats in your barn or enclosed areas where animals are kept on the property?** | ❑ | ❑ | ❑ | ❑ | ❑ |

### Human Illness History/COVID-19 Section [HMNill-19]

**53) What is your current smoking status?** ***(including tobacco, non-tobacco, vape, and e-cig products)***

- ❑ Never a smoker
- ❑ Not currently smoking, but have smoked in the past
    - *Branching logic*: how long has it been since you quit smoking? *Dropdown:[days, weeks, months, years]*
- ❑ Currently smoking
    - *Branching logic:* On average how many cigarettes do you smoke a day?

**54) Have you smoked any products in the last 30 days?** ***(including tobacco, non-tobacco vape, and e-cig products)***

- ❑ No
- ❑ Yes

**54 b) On average how often do you smoke?**

- ❑ Daily

- ❑ A few times per week
- ❑ A few times per month

**54 c) Do you smoke inside the home?**

- ❑ Yes
- ❑ No

**54 d) Have any household members smoked any products in the last 30 days?** *(including tobacco, non-tobacco, vape, and e-cig products)*

- ❑ No
- ❑ Yes → *Branching logic*: Household member identification chart:

| | On average how often have they smoked tobacco products? | Do they smoke inside the home? |
|---|---|---|
| Household members Initials will self-populate | ❑ Daily<br>❑ A few times per week<br>❑ A few times per month | ❑ Yes<br>❑ No |

### Participant Health

**55) Do you have any long-term illnesses or long-term health problems?**

- ❑ Yes
- ❑ No
- ❑ Don't Know

**55 b) If yes please what is/are your long-term illness(es) or long-term health problem(s):**

____________________________________________________________

**56) Have you EVER been told by a healthcare professional that you have any of the following diseases?** *(Select all that apply)*

- ❑ Heart failure
- ❑ Coronary artery disease
- ❑ Cancer
- ❑ Rheumatoid arthritis
- ❑ Asthma
- ❑ COPD
- ❑ Renal insufficiency or chronic kidney disease
- ❑ Immune deficiency
- ❑ Autoimmune disorder
- ❑ Diabetes
  - *Branching logic:*
    - Type 1
      - *branching logic: have you currently or recently (within the last 30 days) received treatment for this diagnosis?* ☐Yes ☐No

- Type 2
  - *branching logic: have you currently or recently (within the last 30 days) received treatment for this diagnosis?* ☐Yes ☐No
- Gestational diabetes
  - *branching logic: have you currently or recently (within the last 30 days) received treatment for this diagnosis?* ☐Yes ☐No
- Don't know the type

- ❑ High blood sugar
- ❑ Hypertension or high blood pressure
- ❑ No, I have never been told I had any of these diseases

*{Branching logic will appear for every disease selected in question 56}*

- *are you currently or have you recently (within the last 30 days) received treatment for this diagnosis?* ☐Yes ☐No

**57) Since January 1st, 2020, has a healthcare professional told you that you have had any of the following diseases?** *(select all that apply)*

- ❑ Flu or Influenza
- ❑ Strep throat *(Streptococcus pyogenes)*
- ❑ Infectious mononucleosis ("Mono")
- ❑ Pneumonia
- ❑ Bronchitis
- ❑ Colitis
- ❑ None of the above

*{Branching logic will appear for every disease selected in question 57}*

- *are you currently under treatment for this diagnosis* ☐ Yes☐ No

**58) Since January 1st, 2020, have you experienced any of the following symptoms** *(select all that apply)***:**

- ❑ Fever
- ❑ Cough
- ❑ Sore throat
- ❑ Chills
- ❑ Shortness of breath
- ❑ Fatigue
- ❑ Headache
- ❑ Chest pain
- ❑ Pneumonia
- ❑ Loss of appetite
- ❑ Diarrhea
- ❑ Loss of sense of taste and/or smell
- ❑ No symptoms
- ❑ Don't Know

**58 b)** *{will appear for each symptom selected}:* **On approximately what date did you start feeling this symptom?**

| *{will appear for each symptom selected from Q58}:* | **On approximately what date did you start feeling this symptom?** |
|---|---|
| **Fever** | ❑ *Calendar date fill-in* |
| **Cough** | ❑ *Calendar date fill-in* |

**58 c)** *{Will appear for each symptom selected}:* **On approximately what date did you stop feeling this symptom?**

| *{will appear for each symptom selected from Q58}:* | **On approximately what date did you stop feeling this symptom?** |
|---|---|
| **Fever** | ❑ *Calendar date fill-in*<br>❑ *I still have this symptom* |
| **Cough** | ❑ *Calendar date fill-in*<br>❑ *I still have this symptom* |

**59)** *{will appear for each symptom selected}:* **Do you believe that when you had this symptom, that you had COVID-19?**

| *{self-populated symptom selected from Q#58}* | **Do you believe that when you had this symptom, that you had COVID-19?** |
|---|---|
| **Fever** | ❑ No, I do not believe that I had COVID-19 when I had this symptom<br>❑ Yes, I believe that I had COVID-19 when I had this symptom<br>○ *Branching logic*: →Why do you believe you had COVID-19 when you had this symptom<br>▪ Yes, I believe that I could have had COVID-19 although I was not tested<br>▪ Yes, I believe I had COVID-19 because a health care provider said I had it |

| | |
|---|---|
| | ▪ Yes, I had a swab or serum test that confirmed I had COVID-19<br>❑ I do not know if I had COVID-19<br>❑ Other (describe): |
| Cough | ❑ No, I do not believe that I had COVID-19 when I had this symptom<br>❑ Yes, I believe that I had COVID-19 when I had this symptom<br>o *Branching logic*: →Why do you believe you had COVID-19 when you had this symptom<br>▪ Yes, I believe that I could have had COVID-19 although I was not tested<br>▪ Yes, I believe I had COVID-19 because a health care provider said I had it<br>▪ Yes, I had a swab or serum test that confirmed I had COVID-19<br>❑ I do not know if I had COVID-19<br>❑ Other (describe): |

**59 b)** *{will only appear for Covid-19 identified symptoms from Q# 59}***: Please check each box next to any medical interventions you have received since developing these COVID-19 related symptoms:** *check all that apply*

**{List of Covid-19 symptoms identified in Q# 59}**

- ❑ Received a COVID-19 diagnostic test
- ❑ Took ibuprofen (Advil), naproxen, aspirin or other NSAID
- ❑ Took acetaminophen (Tylenol)
- ❑ Was prescribed an anti-viral medication
- ❑ Met with a Clinician via telemedicine
- ❑ Visited a clinic, doctor's office, hospital or emergency room for care
- ❑ Admitted to the hospital for care
- ❑ Received oxygen
- ❑ Received sedation
- ❑ Placed on the mechanical ventilator
- ❑ Took or was prescribed other medication
  - b) *Branching logic*: please describe the other mediation you took:__ ____________________________________________
- ❑ Received additional types of medical care or treatment
  - b) Branching logic: please describe the other types of medical care or treatment: ____________________________________________
- ❑ No medical intervention

**59 c)** *{will only appear for COVID-19 identified symptoms from Q# 59 and if pets were identified}* **While experiencing these COVID-19 related symptoms, did you limit your interaction with your pets in any of the following ways?**

**{List of Covid-19 symptoms identified in Q# 59}**

- ❑ I stopped touching my pets and separated myself completely.
- ❑ I did not separate myself completely, but I stopped touching my pets
- ❑ I occasionally touched my pets *Branching logic*→ 59 d, 59 e
- ❑ I did not limit my interactions with my pets *Branching logic*→ 59 d, 59 e

**59 d)** *{will only appear for Covid-19 identified symptoms from Q# 59}* **Did you ever wash your hands or use hand sanitizer BEFORE touching your pet while experiencing these symptoms?**

**{List of Covid-19 symptoms identified in Q# 59}**

- ❑ Always
- ❑ Most of the time
- ❑ Sometimes
- ❑ Never
- ❑ Don't Know

**59 e)** *{will only appear for Covid-19 identified symptoms from Q# 59}* **Did you ever wash your hands or use hand sanitizer AFTER touching your pet while experiencing these symptoms?**

**{List of Covid-19 symptoms identified in Q# 59}**

- ❑ Always
- ❑ Most of the time
- ❑ Sometimes
- ❑ Never
- ❑ Don't know

**60)** *{will only appear if selected working with animals in Q# 27 and for Covid-19 identified symptoms Q# 59}* **While experiencing these symptoms, did you limit interaction with animals associated with your work in any of the following ways?** *For example, if you are a farmer, we are interested in interactions you may have had with farm animals at work. Or if you are a veterinary technician, we are interested in interactions you may have had with animal patients.*

- ❑ I stopped working and had no contact with animals associated with my work
- ❑ I did not isolate myself completely, but I stopped touching animals at work
- ❑ I occasionally touched animals at work *Branching logic*→ 60 b, 60 c
- ❑ I did not limit my interactions with animals at work *Branching logic*→ 60 b, 60 c

**60 b) Did you ever wash your hands or use hand sanitizer BEFORE touching animals at work while experiencing these symptoms?**

**{List of Covid-19 symptoms identified in Q# 59}**

- ❑ Always
- ❑ Most of the time
- ❑ Sometimes
- ❑ Never

- ❑ Don't Know

**60 c) Did you ever wash your hands or use hand sanitizer AFTER touching animals at work while experiencing these symptoms?**
**{List of Covid-19 symptoms identified in Q# 59}**

- ❑ Always
- ❑ Most of the time
- ❑ Sometimes
- ❑ Never
- ❑ Don't Know

### Household Member Health

**61) Do one or more household members have any long-term illnesses or long-term health problems?**

- ❑ Yes
    - *Branching logic*: Please identify those household members
      *{Household member Initial will self-populate}*
      Persons Initials__________
- ❑ No
- ❑ Don't Know

**61 b) If Yes, please describe what is/are their long-term illness(es) or long-term health problem(s):**

________________________________________________________________________________

**62) Have one or more members of your household EVER been diagnosed with or treated for any of the following diseases?** *(select all that apply)*

- ❑ Heart failure
- ❑ Coronary artery disease
- ❑ Cancer
- ❑ Rheumatoid arthritis
- ❑ Hypertension
- ❑ Asthma
- ❑ COPD
- ❑ Renal insufficiency or chronic kidney disease
- ❑ Immune deficiency
- ❑ Autoimmune disorder
- ❑ Diabetes
    - Branching logic:
        - Type 1

- *branching logic: have they currently or recently (within the last 30 days) received treatment for this diagnosis?* ☐Yes ☐No
- Type 2
  - *branching logic: have they currently or recently (within the last 30 days) received treatment for this diagnosis?* ☐Yes ☐No
- Gestational diabetes
  - *branching logic: have they currently or recently (within the last 30 days) received treatment for this diagnosis?* ☐Yes ☐No
- Don't know the type

❑ High blood sugar
❑ Hypertension or high blood pressure

*{Branching logic will appear for every disease selected in question #62}:*

- *Please identify those household members {Household member Initial will self-populate} Persons Initials______*
- *Have they currently or recently (within the last 30 days) received treatment for this diagnosis? ☐Yes ☐No*

**63) Since January 1st, 2020, have one or more members of your household been diagnosed with or treated for any of the following diseases?** *(select all that apply)*

❑ Flu or Influenza
❑ Strep throat *(Streptococcus pyogenes)*
❑ Mono or Infectious mononucleosis
❑ Pneumonia
❑ Bronchitis
❑ Colitis
❑ None of the above

*{Branching logic will appear for every disease selected in question #63}:*

- Please identify those household members *{Household member Initial will self-populate}* Persons Initials________
- *are they **currently** under treatment for this diagnosis* ☐ *Yes*☐ *No*

**64) Since January 1st, 2020, have any household members experienced any of the following symptoms** *check all that apply***:**

❑ Fever
❑ Diarrhea
❑ Cough
❑ Sore throat
❑ Chills
❑ Shortness of breath
❑ Fatigue
❑ Headache
❑ Chest pain

- ❑ Pneumonia
- ❑ Loss of appetite
- ❑ Loss of sense of taste and/or smell
- ❑ No symptoms
- ❑ Don't Know

*{Branching logic will appear for every disease selected in question #64}*:

- ○ Please identify those household members *{Household member Initial will self-populate}*
  Persons Initials___________

**64 b)** *{will appear for each symptom selected}*: **On approximately what date did they START feeling this symptom?** {redcap calendar matrix}

| Household members Initials will self-populate | **On approximately what date did they start feeling this symptom?** **{List of symptoms selected from Q# 64}** |
|---|---|
| Household member A | ❑ *Fever*<br>○ *Branching logic: Calendar date fill-in*<br>❑ Cough<br>○ *Branching logic: Calendar date fill-in*<br>❑ Sore throat<br>○ *Branching logic: Calendar date fill-in*<br>❑ Chills<br>○ *Branching logic: Calendar date fill-in* |
| Household member B | ❑ Chest pain<br>○ *Branching logic: Calendar date fill-in*<br>❑ Headache<br>○ *Branching logic: Calendar date fill-in* |

**64 c)** *{will appear for each symptom selected}*: **On approximately what date did they STOP feeling this symptom?** {redcap calendar matrix}

| Household members Initials will self-populate | **On approximately what date did they stop feeling this symptom?** **{List of symptoms selected from Q# 64}** |
|---|---|

| | |
|---|---|
| Household member A | ❑ *Fever*→*Branching logic:*<br>o *Calendar date fill-in*<br>o *They still have this symptom*<br>❑ Cough→*Branching logic:*<br>o *Calendar date fill-in*<br>o *They still have this symptom*<br>❑ Sore throat-→*Branching logic:*<br>o *Calendar date fill-in*<br>o *They still have this symptom*<br>❑ Chills→*Branching logic:*<br>o *Calendar date fill-in*<br>o *They still have this symptom* |
| Household member B | ❑ Chest pain →*Branching logic:*<br>o *Calendar date fill-in*<br>o *They still have this symptom*<br>❑ Headache→*Branching logic:*<br>o *Calendar date fill-in*<br>o *They still have this symptom* |

**65)** *{will appear for each symptom selected}***: Do you believe that when your household member had this symptom, that they had COVID-19?** *check all that apply*

| Household members Initials will self-populate<br>Household member A | **Do you believe that when this household member had this symptom, that they had COVID-19?** *check all that apply* |
|---|---|
| *{self-populated symptom}*<br><br>*Fever* | ❑ No, I do not believe that they had COVID-19 when they had this symptom<br>❑ Yes, I believe that they had COVID-19 when they had this symptom<br>o *Branching logic*: →Why do you believe they had COVID-19 when they had this symptom?<br>▪ Yes, I believe that they could have had COVID-19 although they were not tested<br>▪ Yes, I believe they had COVID-19 because a health care provider said they had it<br>▪ Yes, they had a swab or serum test that confirmed they had COVID-19<br>❑ I do not know if they had COVID-19<br>Other (describe): |
| Cough | ❑ No, I do not believe that they had COVID-19 when they had this symptom<br>❑ Yes, I believe that they had COVID-19 when they had this symptom |

<table>
<tr><td></td><td>
<ul><li style="list-style-type:circle">Branching logic: →Why do you believe they had COVID-19 when they had this symptom?
<ul><li>Yes, I believe that they could have had COVID-19 although they were not tested</li>
<li>Yes, I believe they had COVID-19 because a health care provider said they had it</li>
<li>Yes, they had a swab or serum test that confirmed they had COVID-19</li></ul></li></ul>
❑ I do not know if they had COVID-19<br>
❑ Other (describe):
</td></tr>
</table>

<table>
<tr><td>Household members Initials will self-populate<br>Household member B</td><td>Do you believe that when this household member had this symptom, that they had COVID-19? check all that apply</td></tr>
<tr><td>{self-populated symptom}<br><br>Chest pain</td><td>
❑ No, I do not believe that they had COVID-19 when they had this symptom<br>
❑ Yes, I believe that they had COVID-19 when they had this symptom
<ul><li style="list-style-type:circle">Branching logic: →Why do you believe they had COVID-19 when they had this symptom?
<ul><li>Yes, I believe that they could have had COVID-19 although they were not tested</li>
<li>Yes, I believe they had COVID-19 because a health care provider said they had it</li>
<li>Yes, they had a swab or serum test that confirmed they had COVID-19</li></ul></li></ul>
❑ I do not know if they had COVID-19<br>
❑ Other (describe):
</td></tr>
<tr><td>Headache</td><td>
❑ No, I do not believe that they had COVID-19 when they had this symptom<br>
❑ Yes, I believe that they had COVID-19 when they had this symptom
<ul><li style="list-style-type:circle">Branching logic: →Why do you believe they had COVID-19 when they had this symptom?
<ul><li>Yes, I believe that they could have had COVID-19 although they were not tested</li>
<li>Yes, I believe they had COVID-19 because a health care provider said they had it</li>
<li>Yes, they had a swab or serum test that confirmed they had COVID-19</li></ul></li></ul>
❑ I do not know if they had COVID-19<br>
❑ Other (describe):
</td></tr>
</table>

**65 b)** *{will only appear for Covid-19 identified symptoms from Q# 65}* **To the best of your ability, please check each box next to any medical interventions other household members received since developing these symptom:** *check all that apply*

| Household members Initials will self-populate | *Covid-19 identified symptoms from Q# 65* | **To the best of your ability, please check each box next to any medical interventions other household members received since developing this symptom {***check all that apply***}** |
|---|---|---|
| Household member A | **Fever/cough** | ❑ Received a COVID-19 diagnostic test<br>❑ Took ibuprofen or other NSAID<br>❑ Took Tylenol<br>❑ Was prescribed an anti-viral<br>❑ Met with a Clinician via telemedicine<br>❑ Visited a clinic, doctor's office, hospital or emergency room for care<br>❑ Admitted to the hospital for care<br>❑ Received oxygen<br>❑ Received sedation<br>❑ Placed on the mechanical ventilator<br>❑ Took or was prescribed other medication<br>b) *Branching logic*: please describe the other mediation they took:<br>________________________________________<br>❑ Received additional types of medical care or treatment<br>b) *Branching logic*: please describe the other types of medical care or treatment:<br>________________________________________<br>❑ No medical intervention |
| Household member B | **Chest pain** | ❑ Received a COVID-19 diagnostic test<br>❑ Took ibuprofen or other NSAID<br>❑ Took Tylenol<br>❑ Was prescribed an anti-viral<br>❑ Met with a Clinician via telemedicine<br>❑ Visited a clinic, doctor's office, hospital or emergency room for care<br>❑ Admitted to the hospital for care<br>❑ Received oxygen<br>❑ Received sedation<br>❑ Placed on the mechanical ventilator<br>❑ Took or was prescribed other medication<br>b) *Branching logic*: please describe the other mediation they took:<br>________________________________________<br>❑ Received additional types of medical care or treatment<br>b) *Branching logic*: please describe the other types of medical care or treatment: |

| | | |
|---|---|---|
| | | ❑ No medical intervention |

**65 c)** *{will only appear for Covid-19 identified symptoms from Q# 65 and pets were identified}*:
**While this household member(s) experienced these symptoms, did they limit interaction with household pets in any of the following ways?**

| Household members Initials will self-populate | *Covid-19 identified symptoms from Q# 65* | **While this household member(s) experienced these symptoms, did they limit interaction with household pets in any of the following ways** |
|---|---|---|
| Household member A | **Fever/cough** | ❑ They stopped touching any pets and completely separated themselves from any pets<br>❑ They did not separate themselves completely, but stopped touching any pets<br>❑ They occasionally touched any pets *Branching logic*→65 d, 65e<br>❑ They did not limit their interactions with any pets *Branching logic*→65 d, 65 e<br>❑ Don't know |
| Household member B | **Chest pain** | ❑ They stopped touching any pets and completely separated themselves from any pets<br>❑ They did not separate themselves completely, but stopped touching any pets<br>❑ They occasionally touched any pets *Branching logic*→65 d, 65 e<br>❑ They did not limit their interactions with any pets *Branching logic*→65 d, 65 e<br>❑ Don't know |

**65 d)** *{will only appear for Covid-19 identified symptoms from Q#65}* **While this household member experienced these symptoms, did they ever wash their hands BEFORE touching any household pets?**

| | Always | Most of the Time | Sometimes | Never | Don't Know |
|---|---|---|---|---|---|

| | | | | | |
|---|---|---|---|---|---|
| Household members Initials will self-populate | ❑ | ❑ | ❑ | ❑ | ❑ |

**65 e) While this household member experienced these symptoms, did they ever wash their hands AFTER touching any household pets?**

| | **Always** | **Most of the Time** | **Sometimes** | **Never** | **Don’t Know** |
|---|---|---|---|---|---|
| Household members Initials will self-populate | ❑ | ❑ | ❑ | ❑ | ❑ |

### COVID-19

**66) Have you ever been told by a healthcare professional that you or any household member may have been infected with COVID-19?** *check all that apply*

- ❑ Yes, myself
- ❑ Yes, one or more members of my household
- ❑ No
- ❑ Don’t know

**67) Have you or any household member been tested for COVID -19?** *check all that apply*

- ❑ Yes, myself
- ❑ Yes, one or more members of my household
    - *Branching logic*: Please identify those household members *{Household member Initial will self-populate*}
      Persons Initials_______________
- ❑ No

**67 b) If Yes, Have you or any household member tested POSITIVE for COVID -19?** ***check all that apply***

- ❑ Yes, myself
- ❑ Yes, one or more members of my household
    - *Branching logic*: Please identify those household members *{Household member Initial will self-populate}*
      Persons Initials_________________
- ❑ No

**67 c) Please identify the type of COVID-19 test that was performed, the date it was done, and the result for ALL COVID -19 TEST you and a household member has taken:**

| | **Nasal Swab** | **Saliva Sample collection** | **Blood Draw** | **Other** |
|---|---|---|---|---|
| The [participant] | **Date:**<br>**Test result:**<br>*Negative, Positive, Inconclusive* | **Date:**<br>**Test result:**<br>*Negative, Positive, Inconclusive* | **Date:**<br>**Test result:**<br>*Negative, Positive, Inconclusive* | **Date:**<br>**Test result:**<br>*Negative, Positive, Inconclusive* |
| Household members Initials will self-populate | **Date:**<br>**Test result:**<br>*Negative, Positive, Inconclusive* | **Date:**<br>**Test result:**<br>*Negative, Positive, Inconclusive* | **Date:**<br>**Test result:**<br>*Negative, Positive, Inconclusive* | **Date:**<br>**Test result:**<br>*Negative, Positive, Inconclusive* |

**68) Have you been sick, or suspected of being sick, with COVID-19 and needed to separate or isolate yourself from people who were not sick?**

- ❑ No, I have not needed to separate or isolate myself from people who were not sick
- ❑ Yes, I had to separate myself or isolate myself from people who were not sick
  - b) *Branching logic:* please provide the start date of this isolation
    Start Date: ____________________________
- ❑ Don't Know

**68 b) Were any of the pets allowed in rooms or areas where you were separating or isolating yourself while you were sick, or suspected of being sick, with COVID-19?**

- ❑ Yes
- ❑ No
- ❑ Don't Know

**68 c) If Yes – how often were pets allowed in these areas while you were sick:** *Please fill out this section for all the pets identified.*

| *{Pet name and ID will be piped-in from first section}* | **Yes, multiple times per day** | **Yes, once per day** | **Yes, a few times a week** | **Yes, once per week** | **Yes, less than weekly** | **Don't Know** |
|---|---|---|---|---|---|---|
| *{Pet_name_1}* | ❑ | ❑ | ❑ | ❑ | ❑ | ❑ |
| *{Pet_name_2}* | ❑ | ❑ | ❑ | ❑ | ❑ | ❑ |
| *{Pet_name_3}* | ❑ | ❑ | ❑ | ❑ | ❑ | ❑ |

**69) Has a household member been sick, or was suspected of being sick, with COVID-19 and needed to separate themselves from people who were not sick?**

- ❑ No household member had needed to separate or isolate themselves from people who were not sick
- ❑ Yes, one or more household members have had to separate themselves or isolate from people who were not sick
  - *Branching logic*: Please identify those household members and their occupation *{Household member Initial will self-populate}* Persons Initials____
  - *Branching logic:* please provide the start date of this isolation
    - Start Date: ___________________________
- ❑ Don't Know

**69 b) Were any of the pets allowed in rooms or areas where a household member was separating or isolating themselves while they were sick, or suspected of being sick, with COVID-19?**

- ❑ Yes
- ❑ No
- ❑ Don't Know

**69 c) If Yes – how often were pets allowed in these areas while they were sick :** *Please fill out this section for all the pets identified.*

| *{Pet name and ID will be piped-in from first section}* | **Yes, multiple times per day** | **Yes, once per day** | **Yes, a few times a week** | **Yes, once per week** | **Yes, less than weekly** | **Don't Know** |
|---|---|---|---|---|---|---|
| *{Pet_name_1}* | ❑ | ❑ | ❑ | ❑ | ❑ | ❑ |
| *{Pet_name_2}* | ❑ | ❑ | ❑ | ❑ | ❑ | ❑ |
| *{Pet_name_3}* | ❑ | ❑ | ❑ | ❑ | ❑ | ❑ |

**70) Have you been exposed, or suspected of being exposed, to COVID-19 and needed to separate yourself from people who were not sick, in order to see if you become sick?** **{***Check all that apply}*

- ❑ No, I have not needed to separate or restrict myself from people who were not sick
- ❑ Yes, I had to separate myself or restrict myself from people who were not sick
  - b) *Branching logic:* please provide the start date of this isolation
    Start Date: _________{ *Calendar date fill-in* }____________________
- ❑ Don't Know

**70 b) Were any of the pets allowed in rooms or areas where you were separating or restricting your movements from other people after you were exposed to, or suspected of being exposed to COVID-19?**

☐ No ☐Yes ☐ Don't Know

**70 c) If yes – how often were pets allowed in these areas where you were separating yourself from other people:** *Please fill out this section for all the pets identified.*

| *{Pet name and ID will be piped-in from first section}* | **Yes, multiple times per day** | **Yes, once per day** | **Yes, a few times a week** | **Yes, once per week** | **Yes, less than weekly** | **Don't Know** |
|---|---|---|---|---|---|---|
| *{Pet_name_1}* | ❑ | ❑ | ❑ | ❑ | ❑ | ❑ |
| *{Pet_name_2}* | ❑ | ❑ | ❑ | ❑ | ❑ | ❑ |
| *{Pet_name_3}* | ❑ | ❑ | ❑ | ❑ | ❑ | ❑ |

**71) Has a household member been exposed, or suspected of being exposed, to COVID-19 and needed to separate themselves from people who were not sick ,in order to see if that household member becomes sick?**

- ❑ No household member needed to separate or restrict themselves from people who were not sick
- ❑ Yes, one or more household members needed to separate or restrict themselves from people who were not sick
  - *Branching logic:* Please identify those household members and their occupation *{Household member Initial will self-populate}* Persons Initials_*(radio button select)*___
  - *Branching logic:* please provide the start date of this isolation
    - Start Date: _________ ____________________
- ❑ Don't Know

**71 b) Were any of the pets allowed in rooms or areas where a household member was separating or restricting their movements from other people after that household member was exposed to, or suspected of being exposed to COVID-19?**

 No Yes  Don't Know

**71 c) If yes, how often were pets allowed in these areas where that household member was separating themselves from other people :** *Please fill out this section for all the pets identified.*

| *{Pet name and ID will be piped-in from first section}* | **Yes, multiple times per day** | **Yes, once per day** | **Yes, a few times a week** | **Yes, once per week** | **Yes, less than weekly** | **Don't Know** |
|---|---|---|---|---|---|---|
| *{Pet_name_1}* | ❑ | ❑ | ❑ | ❑ | ❑ | ❑ |
| *{Pet_name_2}* | ❑ | ❑ | ❑ | ❑ | ❑ | ❑ |
| *{Pet_name_3}* | ❑ | ❑ | ❑ | ❑ | ❑ | ❑ |

**Animal Worker Section [AW-7]**

*{this section will only appear if working with animals was selected above}*

**72) In a previous section of this survey, you identified that you work with animals, or have contact with farm animals. Please estimate the frequency and type of contact you had with the following animals in the last 30 days.**
**Select all that apply:**

- ❑ Rats or mice
  - *Branching logic:* **Select frequency of contact:**
    - ❑ Daily
    - ❑ Weekly
    - ❑ Monthly
    - ❑ Don't know
  - *Branching logic*: **Select average type of contact:**
    - ❑ Close prolonged touch contact,
    - ❑ Intermittent touch contact,
    - ❑ Infrequent touch contact,
    - ❑ No contact
- ❑ Rabbits, hamsters or gerbils, guinea pigs, ferrets,
- ❑ Dogs, cats,
- ❑ Cattle
- ❑ Sheep, goats
- ❑ Swine or pigs
- ❑ Horses
- ❑ Poultry or other fowl
- ❑ Wild mammals
  - *Branching logic*: List species______
- ❑ Wild birds
  - *Branching logic*: List species_____
- ❑ Monkeys

Other____

**73) In your own words, please describe the nature of contact you had with each of the animals listed above in the last 30 days. Be as specific as possible.** (For example: I feed 12 backyard chickens daily and I clean a horse barn housing 3 horses weekly; or I perform surgery on livestock as part of my veterinary practice, etc.)

(text fill-in)

---

**74) Are you a student, staff or faculty at a Veterinary school, laboratory animal program, agriculture program, dairy program, Veterinary technician programs?**

- ❑ Yes
- ❑ No

**74 b) Please identify which school or program:**

*Dropdown selection*

**74 c) Please identify whether you are a student, staff, or faculty at your institution? (please note year of anticipated graduation, or is staff/faculty your department)**

- ❑ DVM/VMD student
  - *Branching logic: anticipated graduation date*
- ❑ Staff
- ❑ Faculty
- ❑ Other___

**75) In the last 30 days, have you worn any of the following items when working with animals?**

| | Always | Most of the Time | Sometimes | Never |
|---|---|---|---|---|
| Protective eyeglasses | ❑ | ❑ | ❑ | ❑ |
| Face mask | ❑ | ❑ | ❑ | ❑ |
| Lab coat | ❑ | ❑ | ❑ | ❑ |
| Goggles or face shield | ❑ | ❑ | ❑ | ❑ |
| Disposable gloves like Nitrile or latex gloves | ❑ | ❑ | ❑ | ❑ |
| Non-disposable gloves like gardening or working gloves | ❑ | ❑ | ❑ | ❑ |
| Scrubs | ❑ | ❑ | ❑ | ❑ |
| Dedicated clothing or other scrubs | ❑ | ❑ | ❑ | ❑ |
| Other protective wear (please specify) | | | | |

**75 b)** *{if face mask is selected}* **Please describe the type of face mask you wear:**

- ❑ Cloth mask
- ❑ Surgical mask
- ❑ N95 mask
- ❑ Respirator

- ❑ Other______

**76) Since January 1st 2020, When working with animals, how often do you wash your hands or use hand sanitizer AFTER interacting with animals?**

- ❑ Always
- ❑ Most of the time
- ❑ Sometimes
- ❑ Never

**77) What percentage of time would you estimate you wash your hands or use hand sanitizer AFTER interacting with animals?**

________________________

**78) In the last 30 days, please tell us about your work-wear routine. How often do you do the following?**

| | Always | Most of the time | Sometimes | Never |
|---|---|---|---|---|
| I change out of my work clothes or scrubs before I leave work | ❑ | ❑ | ❑ | ❑ |
| I change out of my work clothes or scrubs when I get home | ❑ | ❑ | ❑ | ❑ |
| I wash my clothes or scrubs each day after work | ❑ | ❑ | ❑ | ❑ |
| I use an employer-provided laundry service to launder my clothes | ❑ | ❑ | ❑ | ❑ |
| I wear the same work clothes or scrubs multiple days during the week | ❑ | ❑ | ❑ | ❑ |
| I wear the shoes/boots I wear at work when I am in my home | ❑ | ❑ | ❑ | ❑ |

### Pet Health History [PetHlth-27]

**79) Have any of your pets been kenneled, boarded, or attended daycare, since January 1st 2020?**

- ❑ Yes *Branching logic:* →Please select which pets: *{Pet names pipped in/ radio button selections}*
- ❑ No

**79 b) If Yes, has your pet been kenneled, boarded or attended daycare at any point within the past:**

*{Pet name and ID will be piped-in}*

**1) Pet Name & Pet ID** *(radio button selections)*

a) 1 month (30 days) ☐**No** ☐**Yes** ☐ **Don't Know**

b) 3 months (90 days) ☐**No** ☐**Yes** ☐ **Don't Know**

c) 6 months ☐**No** ☐**Yes** ☐ **Don't Know**

d) Year ☐**No** ☐**Yes** ☐ **Don't Know**

**79 c) How many days in total has your pet been kenneled in the last 6 months?**
*{Pet name and ID will be piped-in from first section}*

**Pet Name & Pet ID** *(radio button selections)*
**Number of Days___***Dropdown selection: 1-100*_________

**79 d)** *{will only appear if 30 days was selected}* **In the last 30 days, how often has your pet(s) attend a daycare or boarding kennel?**

- ❑ Daily
- ❑ Twice per week
- ❑ Once per week
- ❑ A few times a month
- ❑ Don't know
- ❑ N/A has not attended daycare/kennel in the last 30 days

**80) Have any of your pets visited a veterinary practice or clinic for any reason since January 1st 2020?** *(Any reason includes routine appointment, hospitalization, nail trim, etc. – this answer should include both outpatient and inpatient visits)*

- ❑ Yes→ Please select which pets: *{Pet names pipped in/ radio button selections}*
- ❑ No

**80 b) If Yes, has your pet visited a veterinary practice or clinic within the past:**
*{Pet name and ID will be piped-in}*
**1) Pet Name & Pet ID** *(radio button selections)*
a) 1 month (30 days) ☐**No** ☐**Yes** ☐**Don't Know**
b) 3 months (90 days) ☐**No** ☐**Yes** ☐**Don't Know**
c) 6 months ☐**No** ☐**Yes** ☐**Don't Know**
d) Year ☐**No** ☐**Yes** ☐**Don't Know**

**80 c) If YES, how many visits to the veterinary practice or clinic have any of your pets made in the past 6 months?**
*{Pet name and ID will be piped-in from first section}*

**1) Pet Name & Pet ID**
**Number of visits________** *Dropdown selection: 1-100*____
**Reason for visit:_________________________**

**81) Have any of your pets been admitted to the veterinary hospital since January 1st, 2020?**

- ❑ Yes→ Please select which pets: *{Pet names pipped in/ radio button selections}*
- ❑ No

**81 b) If Yes, have any of your pets been admitted to the veterinary hospital at any point within the past:**
a) 1 month (30 days) ☐**No** ☐**Yes** ☐**Don't Know**
b) 3 months (90 days) ☐**No** ☐**Yes** ☐**Don't Know**
c) 6 months ☐**No** ☐**Yes** ☐**Don't Know**

d) Year ☐**No** ☐**Yes** ☐**Don't Know**

**81 c) If YES, how many days in total was your pet admitted within the past 6 months?**
*{Pet name and ID will be piped-in from first section}*

**1) Pet Name & Pet ID**
**Number of Days___***Dropdown selection: 1-100***_________**
**Reason for being admitted:__________________________**

**82) Have any of your pets had diarrhea within the past:**
a) 1 month (30 days) ☐**No** ☐**Yes** ☐**Don't Know**
b) 3 months (90 days) ☐**No** ☐**Yes** ☐**Don't Know**
c) 6 months ☐**No** ☐**Yes** ☐**Don't Know**
d) Year ☐**No** ☐**Yes** ☐**Don't Know**

**82 b) If yes, which Pet?***{Pet name and ID will be piped-in from first section}*
**Pet Name & Pet ID** *(radio button selections)*

**83) Have any of your pets had a cough and/or brought up phlegm in the past:**
a) 1 month (30 days) ☐**No** ☐**Yes** ☐ **Don't Know**
b) 3 months (90 days) ☐**No** ☐**Yes** ☐ **Don't Know**
c) 6 months ☐**No** ☐**Yes** ☐ **Don't Know**
d) Year ☐**No** ☐**Yes** ☐ **Don't Know**

**83 b) If yes, which Pet?** *{Pet name and ID will be piped-in from first section}*
**Pet Name & Pet ID** *(radio button selection* **Response:__** (scroll down)**________**

**84) Have any of your pets had wheezing and/or whistling in the chest in the past:**
a) 1 month (30 days) ☐**No** ☐**Yes** ☐ **Don't Know**
b) 3 months (90 days) ☐**No** ☐**Yes** ☐ **Don't Know**
c) 6 months ☐**No** ☐**Yes** ☐ **Don't Know**
d) Year ☐**No** ☐**Yes** ☐ **Don't Know**

**84 b) If yes, which Pet?** *{Pet name and ID will be piped-in from first section}*
**Pet Name & Pet ID** *(radio button selection* **Response:___** (scroll down)**________**

**85) Have any of your pets had sneezing episodes in the past:**
a) 1 month (30 days) ☐**No** ☐**Yes** ☐ **Don't Know**
b) 3 months (90 days) ☐**No** ☐**Yes** ☐ **Don't Know**
c) 6 months ☐**No** ☐**Yes** ☐ **Don't Know**
d) Year ☐**No** ☐**Yes** ☐ **Don't Know**

**85 b) If yes, which Pet?***{Pet name and ID will be piped-in from first section}*
**Pet Name & Pet ID** *(radio button selection* **Response:** (scroll down)**________**

**86) In the past three months, have you noticed any discharge from any of your pet's noses? If so, please note the type of discharge.**

| | **No Discharge** | **Clear** | **Cloudy** | **Bloody** | **Combination** |
|---|---|---|---|---|---|
| **PetName&ID** | ❑ | ❑ | ❑ | ❑ | ❑ |

**87) Have any of your pets been having difficulty breathing *(For example open mouth breathing, or panting in cats, excessive panting, or labored breathing in dogs)* in the past:**
a) 1 month (30 days) ☐**No** ☐**Yes** ☐ **Don't Know**
b) 3 months (90 days) ☐**No** ☐**Yes** ☐ **Don't Know**
c) 6 months ☐**No** ☐**Yes** ☐ **Don't Know**
d) Year ☐**No** ☐**Yes** ☐ **Don't Know**

**87 b) If yes, which Pet?** *{Pet name and ID will be piped-in from first section}*
**Pet Name & Pet ID** *(radio button selection* **Response:** (scroll down)________

**88) Have any of your pets been vaccinated against rabies?**
☐**No**
☐**Yes, vaccinated over a year ago**
☐**Yes, vaccinated in the past year**
☐**N/A (not a mammal)**
☐ **Don't Know**

**88 b) If yes, which Pet?** *{Pet name and ID will be piped-in from first section}*
**Pet Name & Pet ID** *(radio button selection* **Response:** (scroll down)________

**89) Have any of your pet DOGS been vaccinated for influenza (Nobivac Canine Flu H3N8/H3N2 or Vanguard CIV H3N2/H3N8 )?**
☐**No**
☐**Yes, vaccinated over a year ago**
☐**Yes, vaccinated in the past year**
☐**N/A (not a dog)**
☐ **Don't Know**

**89 b) If yes, which Pets?**
*{Pet name and ID will be piped-in from first section}*
**Pet Name & Pet ID** *(radio button selection* **Response:** ___(scroll down)________

**90) Have any of your pet DOGS been vaccinated for Bordetella or Kennel Cough (nasal, oral, or injectable vaccine such as Intratrac, Coughguard, Bronchichine, or Bronchishield)?**
☐**No**
☐**Yes, vaccinated over a year ago**
☐**Yes, vaccinated in the past year**
☐**N/A (not a dog)**
☐ **Don't Know**

**90 b) If yes, which Pets?**

*{Pet name and ID will be piped-in from first section}*
**Pet Name & Pet ID** *(radio button selection)* **Response: ____**(scroll down)**________**

*{Question# 91 will only appear if a pet Cat has been identified}*
**91) Have any or your pet CATS been vaccinated against Feline Infections Peritonitis (FIP), the disease caused by mutation of feline coronavirus (FCoV)?**
**☐No**
**☐Yes, vaccinated over a year ago**
**☐Yes, vaccinated in the past year**
**☐N/A**
**☐ Don't Know**

**91 b) If yes, which Pet?**
*{Pet name and ID will be piped-in from first section}*
**Pet Name & Pet ID** *(radio button selection* **Response:** (scroll down)**________**

**92) Have any of your pets taken oral or injectable glucocorticoids or other immunosuppressive drugs, such as prednisone, or chemotherapeutics like cisplatin, vincristine or doxorubicin, since January 1st, 2020?**
**☐No ☐Yes ☐ Don't Know**

**92 b) If yes, which Pet?**
*{Pet name and ID will be piped-in from first section}*
**1) Pet Name & Pet ID** *(radio button selection)*
**Is this pet currently on these medications?**
**☐No ☐Yes ☐ Don't Know**

**93) Have any of your pets been treated with topical glucocorticoids or other immunosuppressive drugs, such as topical steroids, since January 1st, 2020?**
**☐No ☐Yes ☐ Don't Know**

**93 b) If yes, which Pet?**
*{Pet name and ID will be piped-in from first section}*
**1) Pet Name & Pet ID** *(radio button selection)*
**Is this pet currently on these medications?**
**☐No ☐Yes ☐ Don't Know**

**94) Have any of your pets been treated with ACE Inhibitors, such as Benazepril or Enalapril, since January 1st, 2020?**
**☐No ☐Yes ☐ Don't Know**

**94 b) If yes, which Pet?**
*{Pet name and ID will be piped-in from first section}*
**1) Pet Name & Pet ID** *(radio button selection)*
**Is this pet currently on this medication?**

**☐No ☐Yes ☐ Don't Know**

**95) Has a veterinarian ever diagnosed any of your pets with diabetes mellitus?**
**☐No ☐Yes ☐ Don't Know**

**95 b) If yes, which Pet?**
*{Pet name and ID will be piped-in from first section}*
**Pet Name & Pet ID** *(radio button selection)*
**Is this pet currently receiving treatment?**
**☐No ☐Yes ☐ Don't Know**

**96) Has a veterinarian ever diagnosed any of your pets with cancer or neoplasia?**
**☐No ☐Yes ☐ Don't Know**

**96 b) If yes, which Pet?**
*{Pet name and ID will be piped-in from first section}*
**Pet Name & Pet ID** *(radio button selection)*
**Is this pet currently receiving treatment?**
**☐No ☐Yes ☐ Don't Know**

**97) Has a veterinarian ever diagnosed any of your pets with Heart disease (*example, heart worm, heart murmurs, afib)*?**
**☐No ☐Yes ☐ Don't Know**

**97 b) If yes, which Pet?**
*{Pet name and ID will be piped-in from first section}*
**Pet Name & Pet ID** *(radio button selection)*
**Is this pet currently receiving treatment?**
**☐No ☐Yes ☐ Don't Know**

**98) Since January 1$^{st}$ 2020, has your pet been on heartworm monthly preventative medications? (example: Heartgard)**
**☐No ☐Yes ☐ Don't Know**

**98 b) If yes, which Pet?**
*{Pet name and ID will be piped-in from first section}*
**Pet Name & Pet ID** *(radio button selection)*

**99) Has any of your pets ever been diagnosed with immunosuppression or a disease that can produce immunosuppression, such as [CATS] feline AIDS (FIV) or feline leukemia (FeLV) or [DOGS] Cushings disease?**
**☐No ☐Yes ☐ Don't Know**

**99 b) If yes, which Pet?**
*{Pet name and ID will be piped-in from first section}*
**Pet Name & Pet ID** *(radio button selection)*

**99 c) Is this pet currently receiving treatment?**
**☐No ☐Yes ☐ Don't Know**

**100) Have any of your CATS been diagnosed with upper respiratory tract infections such as Feline Calicivirus, Chlamydia felis or Feline herpesvirus (FHV-1) in the past?**
a) 1 month (30 days) **☐No ☐Yes ☐ Don't Know**
b) 3 months (90 days) **☐No ☐Yes ☐ Don't Know**
c) 6 months **☐No ☐Yes ☐ Don't Know**
d) Year **☐No ☐Yes ☐ Don't Know**

**100 b) If yes, which Pet?**
*{Pet name and ID will be piped-in from first section}*
**Pet Name & Pet ID** *(radio button selection)* **Response: ____**(scroll down)**________**

**101) Has a veterinarian diagnosed your any of your pets with influenza in the past:**
a) 1 month (30 days) **☐No ☐Yes ☐ Don't Know**
b) 3 months (90 days) **☐No ☐Yes ☐ Don't Know**
c) 6 months **☐No ☐Yes ☐ Don't Know**
d) Year **☐No ☐Yes ☐ Don't Know**

**101 b) If yes, which Pet?**
*{Pet name and ID will be piped-in from first section}*
**Pet Name & Pet ID** *(radio button selection)* **Response: ____**(scroll down)**________**

**102) Have you been told by a veterinarian that any of your pets could have Bordetella or Kennel Cough in the past:**
a) 1 month (30 days) **☐No ☐Yes ☐ Don't Know**
b) 3 months (90 days) **☐No ☐Yes ☐ Don't Know**
c) 6 months **☐No ☐Yes ☐ Don't Know**
d) Year **☐No ☐Yes ☐ Don't Know**

**102 b) If yes, which Pet?**
*{Pet name and ID will be piped-in from first section}*
**Pet Name & Pet ID** *(radio button selection)* **Response: ___**(scroll down)**________**

**103) Is your pet currently taking oral or injectable antibiotics?** *i.e. antibiotic by mouth or by injection.*
**☐No**
**☐Yes**
**☐ Don't Know**

**103 b) If yes, which Pet?**
*{Pet name and ID will be piped-in from first section}*
**Pet Name & Pet ID** *(radio button selection)*

**104) Has your pet taken oral or injectable antibiotics. in the past:**

b) 1 month (30days) ☐**No** ☐**Yes** ☐ **Don't Know**
c) 3 months (90 days) ☐**No** ☐**Yes** ☐ **Don't Know**
d) 6 months ☐**No** ☐**Yes** ☐ **Don't Know**
e) Year ☐**No** ☐**Yes** ☐ **Don't Know**

**104 b) IF YES to current, 4 weeks, or 6 months:**

How many courses of antibiotics did your pet take in the last six months?
Please list all antibiotics taken in the last six months by name, mm/yy, approx.. # days of tx, & route of administration (oral, SQ, IM, IV etc.):

| Name of Antibiotic (approximate date of treatment if known) |
|---|

Thank you for taking this survey. With your consent we would like to be able to contact you in the future for follow up questions. By selecting "yes" you will be sent to a different URL where you can enter your contact information.

**105) Future Contact:**

- ❑ Yes
- ❑ No
