## Supplementary material for "A standardized instrument quantifying risk factors associated with bi-directional transmission of SARS-CoV-2 and other zoonotic pathogens: The COVID-19 Human-Animal Interactions Survey (CHAIS)": Abridged CHAIS instrument (A-CHAIS)

V 1.0 Abridged CHAIS (A-CHAIS)

| |
|---|
| COVID-19 Study ID: |
| Today's Date |
| Interviewer Initials |

*NOTE: This instrument has been populated and is ready to use through RedCap. Please contact the corresponding author Dr. Kaitlin B. Waite DVM MPH at to receive a copy of this instrument. Open online access through RedCap will be made available in the future. Any text below in red represents code instructions in RedCap.*

### Household Demographics [DEM-11]

**1) What is your date of birth?**

**2) What is your current height?**

**3) What is your current weight?**

**4) What is your gender?**

- ❑ Female
- ❑ Male
- ❑ Transgender
- ❑ Nonbinary
- ❑ Other________________
- ❑ Do not wish to answer

**5) What is your race?** *Select all that apply*

- ❑ American Indian or Alaska Native
- ❑ Asian
- ❑ Black or African American
- ❑ Native Hawaiian or Other Pacific Islander
- ❑ White
- ❑ Other__________
- ❑ Don't Know
- ❑ Do not wish to answer

**6) What is your zip code/postal code?**

**7) What type of dwelling do you live in?**

- ❑ Single Family House
- ❑ Townhome
- ❑ Apartment
    - *Branching logic:*
    - How many floors are in your Building?
        - *Dropdown: 2-300, Don't know*

- Please estimate how many units/apartments are in your Building*:*
  - *Dropdown: 2-300, Don't know*

❑ Condominium
- *Branching logic:*
- How many floors are in your building?
  - *Dropdown: 2-300, Don't know*
- Please estimate how many units are in your building*:*
  - *Dropdown: 2-300, Don't know*

❑ Other

**8) What is the approximate size of your home in square footage?**

❑ Less than 500 square feet
❑ 500-1000 square feet
❑ 1001-3000 square feet
❑ 3001 or more square feet
❑ Don't know

**9) How many bedrooms and bathrooms are in your home?** *(if you live in a studio apartment, select "0" for number of bedrooms)*

- Bedrooms _____*{Dropdown: 0-20}*_________
- Bathrooms ____*{Dropdown: 0-20}* ________

**10) How many people, including yourself, live in your household?** *Please enter the number of household members including yourself.*_________ *{Dropdown: 1-20}* ___________

**11) Below, please provide information for up to 8 people currently living with you in your house.**

| Persons ID | Persons Initials | Age in yrs | Sex | Relationship to you |
|---|---|---|---|---|
| *{self-populated hidden field}* | *{free text}* | *{dropdown: <0-100}* | ❑ Female<br>❑ Male<br>❑ Transgender<br>❑ Nonbinary<br>❑ Other<br>❑ Do not wish to answer | ❑ Mother<br>❑ Father<br>❑ Sibling<br>❑ Child<br>❑ Spouse/Partner<br>❑ Other: |

### Pet Demographics and Behaviors [PetDEM-6]

**12) Do you have any pet animals currently living *inside* your home?**

❑ **Yes**
❑ **No**

**12 b) How many total pets do you currently have *inside* your house?**

*By pet, we are referring to an animal, kept primarily for a person's company or entertainment rather than as livestock or a laboratory animal.*

*Note: all animals should be counted individually, except in the event of a group of animals living together in an enclosure, for example, multiple fish in a tank should be grouped and identified as 1 fish tank or multiple hamsters in a cage should be grouped as 1 cage of hamsters.*

- ❑ *{Dropdown scroll 1-50}*

**13) What are your pets NAMES, SPECIES, BREED, SEX, year of birth ,and approximate AGE?**

*Please identify up to 10 of the pets currently living* ***primarily in your house****. Please record your pets in the following order of priority:*

- *1) Companion animal [example cats and dogs]*
- *2) Small companion animals [example rabbits, mink, ferrets]*
- *3) Rodents and Pocket Pet(s) [example: hamsters, gerbils, pet mice, pet rats, guinea pigs]*
- *4) Pet birds [example cockatiel, canaries]*
- *5) Farm animals kept indoors [potbellied pigs, chickens]*
- *6) Other [example water turtles or other reptiles, fish tanks]*

*Note: all animals should be listed individually, except in the event of a group of animals living together in an enclosure, for example multiple fish in a tank should be grouped and identified as 1 fish tank or multiple hamsters in a cage should be grouped as 1 cage of hamsters.*

| Pet Study ID | PETs NAME | Species | BREED | SEX | SEX Alt | Age | How long have you had this pet? |
|---|---|---|---|---|---|---|---|
| *{self-populated hidden field}* | | | | ☐ **Male**<br>☐**Female**<br>☐ **Don't know** | ☐**Neutered**<br>☐ **Spayed**<br>☐ **Don't know**<br>☐**N/A** | ❑ *0-2yrs* *{Branching logic}:* **Age in months** *{dropdown: 1-24mo}*<br>❑ *3-6 yrs*<br>❑ *7-13yrs*<br>❑ *14-100yrs* | |

**14) Do any pet animals spend time <u>outside</u> the home?** *check all that apply*

☐**Yes** (100% Outdoor pet)

☐ **Yes** *(indoor/outdoor: pet spends 30% of their time in either location)*
☐ **No** *(100% indoor pet except for bathroom breaks and leashed/unleashed accompanied walks)*
☐ **Don't Know**

**15) In the last 30 days, how many hours per day has your pet spent outside?** *check all that apply*

| *{Pet name and ID will be piped-in from the first section}* | **100% indoor pet (never outside)** | **Less than 1 hr/ day** | **2-4 hrs /day** | **5-7 hrs/day** | **More than 8 hrs/day** | **100% outdoor pet** |
|---|---|---|---|---|---|---|
| Pet A | ❑ | ❑ | ❑ | ❑ | ❑ | ❑ |
| Pet B | ❑ | ❑ | ❑ | ❑ | ❑ | ❑ |

**16) How would you describe your pet's primary environment with relation to where you live?**

❑ Urban
❑ Suburban
❑ Rural

**17) Do any of your pets currently have direct contact with any of the following animals outside the household?** *Direct contact means touching.*

| *{Pet name and ID will be piped-in from the first section}* | **No Contact** | **Less than once/Month** | **Several times/Month** | **Daily Contact** |
|---|---|---|---|---|
| **a)** Domestic Dog | ❑ | ❑ | ❑ | ❑ |
| **b)** Stray Dog | ❑ | ❑ | ❑ | ❑ |
| **c)** Domestic Cat | ❑ | ❑ | ❑ | ❑ |
| **d)** Feral Cat | ❑ | ❑ | ❑ | ❑ |
| **e)** Wild bird | ❑ | ❑ | ❑ | ❑ |
| **f)** Wild rodent (squirrels, mouse, rat, etc.) | ❑ | ❑ | ❑ | ❑ |
| **g)** Wild Mammals: (bunnies, raccoons, skunks) | ❑ | ❑ | ❑ | ❑ |
| **h)** Horse | ❑ | ❑ | ❑ | ❑ |
| **i)** Pig | ❑ | ❑ | ❑ | ❑ |
| **j)** Cow | ❑ | ❑ | ❑ | ❑ |
| **k)** Chickens or turkeys | ❑ | ❑ | ❑ | ❑ |
| **l)** Sheep | ❑ | ❑ | ❑ | ❑ |

**Other animals (write in): ______________________________________**

*Click here to add another pet that has direct contact with animals outside the home*
*[Another animal contact chart will be made available in RedCap]*

### Occupation Section [Occ-3]

**18) Are you currently employed?**
- ❑ Yes
- ❑ No

**19) In your occupation, do you work with animals?**
- ❑ Yes
- ❑ No *Branching logic: Skip to question 19d*

**19 b) If Yes, what is your occupation?** *Please select all occupation categories that apply to your job***.**
- ❑ Animal breeder
- ❑ Animal trainer
- ❑ Animal control worker
- ❑ Animal scientist or animal researcher
- ❑ Farmer, rancher, farm-worker, or other agricultural worker
- ❑ Non-farm animal caretaker (boarding kennel, doggy daycare, or shelter worker, groomer, etc.)
- ❑ Laboratory animal caretaker
- ❑ Veterinarian
- ❑ Veterinary technologist, technician, or assistant
- ❑ Veterinary student
- ❑ Zoologist or zoo worker
- ❑ Wildlife biologist or wildlife worker
- ❑ Involved in livestock processing for food or fiber production
- ❑ Other_____

**19 c) Please list the most common animals you work with.** *For example: "horses, pigs, sheep, goats"* ____________________________________________________

**19 d) If No, what is your occupation?** _______________________________

**20) In the last 30 days, if your regular workplace is outside your home, <u>on average</u>, how many hours per week have you worked in your regular workplace?**
- ❑ 0 hours
- ❑ 1-10 hours
- ❑ 11-20 hours

- ❑ 21-30 hours
- ❑ 31-40 hours
- ❑ >40 hours
- ❑ N/A (regular workplace not outside the home)

### Travel and Activities Section [Travel-3]

**21) Since January 1st, 2020, have you traveled to another U.S. state other than the one you live in?**

- ❑ Yes, I have traveled to another U.S. state
    - *Branching logic:* Since January 1st, 2020 how many times have you traveled to another U.S state?
- ❑ No, I have not traveled to another U.S. state
- ❑ Don't Know

**21 b) Did your pet(s) travel with you on any of these trips?**

- ❑ Yes → *Branching logic: Please select which pet*
- ❑ No

**22) Since January 1st, 2020, have any household members traveled to another U.S. state other than the one they live in?**

- ❑ Yes, one or more household members have traveled to another U.S. state
- ❑ No household members have traveled to another U.S. state
- ❑ Don't Know

**22 b) If Yes, Did your pet(s) travel with them?**

- ❑ Yes → *Branching logic: Please select which pet*
- ❑ No

**23) In the last 30 days, while not at work, how often have you worn the following personal protective equipment when outside your home?** *Please be as truthful as possible. Remember: this survey is confidential and this question will help us better understand precautionary behaviors during COVID-19. (select all that apply)*

| | Every time I leave the house | Most of the time | Sometimes | Almost never | Never |
|---|---|---|---|---|---|
| **Disposable gloves like Nitrile or latex gloves** | ❑ | ❑ | ❑ | ❑ | ❑ |
| **Non-disposable gloves like gardening or working gloves** | ❑ | ❑ | ❑ | ❑ | ❑ |

| | | | | | |
|---|---|---|---|---|---|
| **Medical grade face masks** | ❑ | ❑ | ❑ | ❑ | ❑ |
| **Other face covering like cloth mask** | ❑ | ❑ | ❑ | ❑ | ❑ |
| **Face shield** | ❑ | ❑ | ❑ | ❑ | ❑ |
| **Other protective wear (please specify)** | | | | | |

**23 b)** *{if medical grade face mask is selected}* **Please describe the type of face mask you wear:**
❑ Surgical mask
❑ N95 or KN95 mask
❑ Respirator
❑ Other_____

### Human-Animal Interaction Section [HAI-9]

**24) Who in your household is most involved in the care of** *{pet name self-populated} {drop down list of household members identified}*

| **24 b) Since January 1st 2020, how often has** *{person identified in question 24-self-generated}* **performed the following roles with/for** *{pet name self-populated }* | **Daily** | **Weekly** | **Monthly** | **Never** |
|---|---|---|---|---|
| Fill food and water | ❑ | ❑ | ❑ | ❑ |
| Hold in arms, lay with, or cuddle | ❑ | ❑ | ❑ | ❑ |
| Given medication (when needed) | ❑ | ❑ | ❑ | ❑ |
| Clean their bedding | ❑ | ❑ | ❑ | ❑ |
| Clean their litter *{only appear if cat}* | ❑ | ❑ | ❑ | ❑ |
| Take outdoors for exercise | ❑ | ❑ | ❑ | ❑ |
| Throw a ball, frisbee, or other toys with your dog when playing "fetch" | ❑ | ❑ | ❑ | ❑ |

| | | | | |
|---|---|---|---|---|
| Play with toys with your cat *{only appear if cat}* | ❑ | ❑ | ❑ | ❑ |
| Play with toys with this pet | ❑ | ❑ | ❑ | ❑ |

*{Question 24 and 24b will repeat and appear for every pet identified in the beginning of the survey}*

**For the following questions regarding intensity of pet contact, please use the following interaction intensity scale:**

| Score | Interaction Intensity |
|---|---|
| 0 | No Contact |
| 1 | Light, gentle petting with hand only |
| 2 | Slightly more assertive petting with hands, allowing pet on lap, holding pet in hand(s) |
| 3 | Heavy-handed petting with hands, hugging the pet, bringing pet close to face |

**25) Please describe the intensity, on average, of pet contact from interactions with humans within the household for** *{pet name self-populated}*

| *Pet Name & Pet ID: pipped in form first section* | **0** | **1** | **2** | **3** |
|---|---|---|---|---|
| You *{The participant}* | ☐ | ☐ | ☐ | ☐ |
| Household member {Household members Initials will self-populate} | ☐ | ☐ | ☐ | ☐ |

*{Question 25 will repeat and appear for every pet identified in the beginning of the survey}*

**26) In the last 30 days, have you, or a household member, kissed any of your pets on the mouth, lips, nose, face, or beak?**
- ❑ Yes
- ❑ No
- ❑ Don’t Know

**26 b) In the last 30 days, have you, or a household member, let any pets touch your face, or a household members face, with pets mouth, lips, nose, face, or beak?**

- ❑ Yes
- ❑ No
- ❑ Don’t Know

**26 c) In the last 30 days, has any pet licked you, or a household member, in the face?**
- ❑ Yes
- ❑ No
- ❑ Don't Know

**26 d) Which pet(s) have you, or a member in your household, kissed on the mouth, lips, nose, face, beak or been touched or licked on the face by?**
*{Check box select of all pets identified}*

**26 e) Who in your household, and at what frequency, kisses** *{pet name}* **on the mouth, lips, nose, face, beak or lets** *{pet name}* **touch their face with its mouth, lips, face, beak or lets** *{pet name}* **lick their face?**

| | **Multiple times per day** | **Once per day** | **A few times a week** | **Once per week** | **A few times in the last 30 days** | **Once in the last 30 days** | **N/A** |
|---|---|---|---|---|---|---|---|
| You *{The participant}* | ❑ | ❑ | ❑ | ❑ | ❑ | ❑ | ❑ |
| Household member {Household members Initials will self-populate} | ❑ | ❑ | ❑ | ❑ | ❑ | ❑ | ❑ |

*{26e will appear for every pet selected on 26d}*

**27) Do you, or any members of your household, allow any of your pets to lick your/their hands?**
- ❑ Yes
- ❑ No
- ❑ Don't Know

**27 b) Which pet(s) lick your, or a household member, hands?** *Select all that apply*
*{Check box select of all pets identified}*

**27 c) Who in your household and at what frequency does** *{pet name}* **lick your, or a household members hands?**

| | **Multiple times per day** | **Once per day** | **A few times a week** | **Once per week** | **A few times in the last 30 days** | **Once in the last 30 days** | **N/A** | **Don't know** |
|---|---|---|---|---|---|---|---|---|
| You *{The participant}* | ❑ | ❑ | ❑ | ❑ | ❑ | ❑ | ❑ | ❑ |

| | | | | | | | | |
|---|---|---|---|---|---|---|---|---|
| Household member {Household members Initials will self-populate} | ❑ | ❑ | ❑ | ❑ | ❑ | ❑ | ❑ | ❑ |

*{27c will appear for every pet selected on 27b}*

**28) Do any of your pets regularly sleep with you or any household members?** *(for example, on the bed, or on the couch, or in a chair)*

- ❑ Yes:→ *Branching logic*: go to 28b
- ❑ No
- ❑ Don't Know

**28 b) Which pets regularly sleep with you or any household members?** *Select all that apply* *{Check box select of all pets identified}*

**28 c) Who in your household does** *{pet name}* **sleep with, and at what frequency?**

### General Farm Animal Contact

**32) Since January 1st, 2020, have you had routine close contact with farm animals?**

- ❑ Yes →*will be shown question block for animal professionals*
- ❑ No
- ❑ Don't Know

**32 b) If yes, how would you best describe your interaction with farm animals?** *Select all that apply*

- ❑ I work in the veterinary medical field and treat farm animals
- ❑ I work on a farm or own farm animals
- ❑ I have other routine interactions with farm animals (please describe)_____________

| **Human Illness History/COVID-19 Section [HMNill-12]** |
|---|

**33) Have you smoked any products in the last 30 days?** ***(including tobacco, non-tobacco, vape, and e-cig products)***

- ❑ No
- ❑ Yes

**33 d) Have any household members smoked any products in the last 30 days?** *(including tobacco, non-tobacco, vape, and e-cig products)*

- ❑ No
- ❑ Yes

### Participant Health

**34) Since January 1st, 2020, have you experienced any of the following symptoms** *(select all that apply)***:**

- ❑ Fever
- ❑ Cough
- ❑ Sore throat
- ❑ Chills
- ❑ Shortness of breath
- ❑ Fatigue
- ❑ Headache
- ❑ Chest pain
- ❑ Pneumonia
- ❑ Loss of appetite
- ❑ Diarrhea
- ❑ Loss of sense of taste and/or smell
- ❑ No symptoms
- ❑ Don't Know

**35)** *{will appear for each symptom selected}***: Do you believe that when you had this symptom, that you had COVID-19?**

| **[self-populated symptom selected from Q#34** | **Do you believe that when you had this symptom, that you had COVID-19?** |
|---|---|
| **Fever** | ❑ No, I do not believe that I had COVID-19 when I had this symptom<br>❑ Yes, I believe that I had COVID-19 when I had this symptom<br>○ *Branching logic*: →Why do you believe you had COVID-19 when you had this symptom<br>▪ Yes, I believe that I could have had COVID-19 although I was not tested<br>▪ Yes, I believe I had COVID-19 because a health care provider said I had it<br>▪ Yes, I had a swab or serum test that confirmed I had COVID-19<br>❑ I do not know if I had COVID-19<br>❑ Other (describe): |

| | |
|---|---|
| Cough | ❑ No, I do not believe that I had COVID-19 when I had this symptom<br>❑ Yes, I believe that I had COVID-19 when I had this symptom<br>○ *Branching logic*: →Why do you believe you had COVID-19 when you had this symptom<br>▪ Yes, I believe that I could have had COVID-19 although I was not tested<br>▪ Yes, I believe I had COVID-19 because a health care provider said I had it<br>▪ Yes, I had a swab or serum test that confirmed I had COVID-19<br>❑ I do not know if I had COVID-19<br>❑ Other (describe): |

sympcovidyes_fever

**35 b)** *{will only appear for COVID-19 identified symptoms from Q#34 and if pets were identified}* **While experiencing these COVID-19 related symptoms, did you limit your interaction with your pets in any of the following ways?**

**{List of Covid-19 symptoms identified in Q#34}**

- ❑ I stopped touching my pets and separated myself completely.
- ❑ I did not separate myself completely, but I stopped touching my pets
- ❑ I occasionally touched my pets *Branching logic*→35 d, 35 e
- ❑ I did not limit my interactions with my pets *Branching logic*→35 d, 35 e

**35 c)** **{***will only appear for Covid-19 identified symptoms from Q#34}* **Did you ever wash your hands or use hand sanitizer BEFORE touching your pet while experiencing these symptoms?**

**{List of Covid-19 symptoms identified in Q#34}**

- ❑ Always
- ❑ Most of the time
- ❑ Sometimes
- ❑ Never
- ❑ Don't Know

**35 d)** **{***will only appear for Covid-19 identified symptoms from Q#34}* **Did you ever wash your hands or use hand sanitizer AFTER touching your pet while experiencing these symptoms?**

**{List of Covid-19 symptoms identified in Q#34}**

- ❑ Always
- ❑ Most of the time
- ❑ Sometimes
- ❑ Never
- ❑ Don't know

**36)** *{will only appear if selected working with animals in Q# 19 and for Covid-19 identified symptoms Q# 34 }* **While experiencing these symptoms, did you limit interaction with animals associated with your work in any of the following ways?** *For example, if you are a farmer, we are interested in interactions you may have had with farm animals. Or if you are a veterinary technician, we are interested in interactions you may have had with animal patients*.

- ❑ I stopped working and had no contact with animals associated with my work
- ❑ I did not isolate myself completely, but I stopped touching animals at work
- ❑ I occasionally touched animals at work *Branching logic*→ 36 b, 36 c
- ❑ I did not limit my interactions with animals at work *Branching logic*→ 36 b, 36 c

**36 b) Did you ever wash your hands or use hand sanitizer BEFORE touching animals at work while experiencing these symptoms?**
**{List of Covid-19 symptoms identified in Q# 34}**

- ❑ Always
- ❑ Most of the time
- ❑ Sometimes
- ❑ Never
- ❑ Don't Know

**36 c) Did you ever wash your hands or use hand sanitizer AFTER touching animals at work while experiencing these symptoms?**
**{List of Covid-19 symptoms identified in Q# 34}**

- ❑ Always
- ❑ Most of the time
- ❑ Sometimes
- ❑ Never
- ❑ Don't Know

### Household Member Health

**37) Since January 1st 2020, have any household members experienced any of the following symptoms** *check all that apply*:

- ❑ Fever
- ❑ Diarrhea
- ❑ Cough
- ❑ Sore throat
- ❑ Chills
- ❑ Shortness of breath
- ❑ Fatigue
- ❑ Headache
- ❑ Chest pain
- ❑ Pneumonia
- ❑ Loss of appetite
- ❑ Loss of sense of taste and/or smell

- ❑ No symptoms
- ❑ Don't Know

*{Branching logic will appear for every symptom selected in question# 37}*

- ○ Please identify those household members *{Household member Initial will self-populate}* Persons Initials_________

**38)** *{will appear for each symptom selected}*: **Do you believe that when your household member had this symptom, that they had COVID-19?** *check all that apply*

**61) Future Contact:**
- ❑ Yes
- ❑ No
